# The dynamic impact of location and resection on the glioma CSF proteome

**DOI:** 10.1101/2024.05.15.24307463

**Authors:** Cecile Riviere-Cazaux, Christopher J. Graser, Arthur E. Warrington, Matthew D. Hoplin, Katherine M. Andersen, Noor Malik, Elizabeth A. Palmer, Lucas P. Carlstrom, Surendra Dasari, Amanda Munoz-Casabella, Samar Ikram, Keyvan Ghadimi, Benjamin T. Himes, Ignacio Jusue-Torres, Jann N. Sarkaria, Fredric B. Meyer, Jamie J. Van Gompel, Sani H. Kizilbash, Ugur Sener, Franziska Michor, Jian L. Campian, Ian F. Parney, Terry C. Burns

## Abstract

While serial sampling of glioma tissue is rarely performed prior to recurrence, cerebrospinal fluid (CSF) is an underutilized longitudinal source of candidate glioma biomarkers for understanding therapeutic impacts. However, the impact of key variables to consider in longitudinal CSF samples, including anatomical location and post-surgical changes, remains unknown. To that end, pre- versus post-resection intracranial CSF samples were obtained at early (1-16 days; n=20) or delayed (86-153 days; n=11) timepoints for patients with glioma. Paired lumbar-versus-intracranial glioma CSF samples were also obtained (n=14). Using aptamer-based proteomics, we identify significant differences in the CSF proteome between lumbar, subarachnoid, and ventricular CSF. Our analysis of serial intracranial CSF samples suggests the early potential for disease monitoring and evaluation of pharmacodynamic impact of targeted therapies. Importantly, we found that resection had a significant, evolving longitudinal impact on the CSF proteome. Proteomic data are provided with individual clinical annotations as a resource for the field.

**One Sentence Summary:** Glioma cerebrospinal fluid (CSF) accessed intra-operatively and longitudinally via devices can reveal impacts of treatment and anatomical location.

## INTRODUCTION

Gliomas are primary brain tumors that inevitably recur despite maximal safe surgical resection and aggressive chemoradiation (*1*). No FDA-approved drug has improved survival for the most common of these tumors, glioblastoma (GBM), since temozolomide (TMZ) in 2005 (*2*). MRIs are standard-of-care for disease monitoring, but remain subject to confounders such as pseudoprogression (*3–7*). Although new techniques are emerging for measuring tumor burden and correlation to glioma biology (*8*), standard MRI sequences do not provide data regarding the biological impacts of treatment. Liquid biopsies may serve as a source of monitoring and pharmacodynamic biomarkers (*9–12*). However, the blood-brain barrier limits passage of CNS-derived analytes (*13*), including glioma biomarkers (*8, 14–18*). In contrast, cerebrospinal fluid (CSF) is a proximal fluid that is an abundant biomarker source for brain tumors(*19–21*).

We and others have evaluated CSF for candidate brain tumor biomarkers such as cell-free DNA (*11, 22–26*), and 2-hydroxyglutarate for IDH-mutant gliomas (*27–30*). The glioma CSF proteome remains relatively underexplored, particularly for longitudinal monitoring of the impact of standard-of-care or experimental therapies (*31*). Since CSF is typically protein-deficient, signal to noise for assessing disease burden and biological impacts of therapy may be superior to plasma. A small number of studies have evaluated the glioma CSF proteome (*32–36*), most often at a single timepoint. However, prior work has described a significant difference between the lumbar and intracranial CSF proteome(*37*) that could impact biomarker identification. Indeed, optimal sensitivity for disease monitoring is obtained from biofluids in contact to tissue (*38, 39*) that can be obtained at the time of surgery or longitudinally via CSF access devices.

Toward our goal of establishing an early understanding of variables impacting the longitudinal glioma CSF proteome that would be of relevance for longitudinal disease monitoring, we performed aptamer-based proteomic analyses utilizing the Somalogic platform (*40, 41*). Of multiple variables that could be evaluated, the impact of anatomical location and of resection were prioritized given their relevance to serial CSF acquisition, namely 1) longitudinal CSF may be sampled from different anatomical locations prior to versus after resection, such as the subarachnoid space upon dural opening and the resection cavity after resection, and 2) most patients with gliomas undergo glioma resection as part of standard-of-care treatment, the longitudinal impacts of which may be relevant for comparison to subsequently administered systemic therapies.

To answer these questions, 146 CSF samples from 74 patients, 71 of whom had grade 2-4 astrocytomas or grade 2-3 oligodendrogliomas, were obtained, spanning different CSF compartments at multiple timepoints, including prior to and after resection. These data include 1) intracranial samples from the subarachnoid space or ventricular system, including both primary or recurrent gliomas, 2) paired lumbar and intracranial samples from patients with low and high-grade gliomas, and 3) longitudinal glioma CSF from CSF access devices, such as Ommaya reservoirs (NCT04692337) (*27*), to evaluate the impact of resection and targeted drugs. We found a significant impact of anatomical location of acquisition on the CSF proteome of gliomas, in addition to a dynamic, evolving, and persistent impact of resection months after surgery. Moreover, the glioma CSF proteome was highly enriched for plasma-derived proteins, suggestive of the impact of blood-brain barrier disruption. All CSF proteomic data and sample annotations are available for each patient included in this study to enable the neuro-oncology community to query these patients’ data for biomarker and biology discovery and validation.

## RESULTS

### Impact of anatomical location on the CSF proteome

Intracranial CSF can be obtained from multiple intracranial locations, including the ventricular system and subarachnoid space. To determine the proteomic impact of intracranial sampling location, we compared CSF from the ventricular system versus subarachnoid space in our most well-powered cohort, patients with GBMs, by performing separate Mann-Whitney U tests for all proteins in our panel while using the Benjamini-Hochberg method to correct for multiple hypothesis testing. We found that 1,033 aptamers had a higher signal in subarachnoid CSF, while 35 had elevated signals in ventricular CSF (FC≥1.5, adjusted p-value≤0.05, **Figure 1Ai**; **Table 1**; full list in **Supplementary Table 1**). Of note, CAPSL was one of the most significant ventricular-associated proteins, consistent with its known predominance in ependymal cells in the ventricle(*42*). Functional protein network analysis suggested enrichment for metabolism-associated pathways in the subarachnoid GBM CSF samples, while ventricular CSF samples were enriched for developmental and immune processes (**Supplementary Table 2).** To evaluate the reproducibility of this subarachnoid versus ventricular signature, the first 10 ventricular and 13 subarachnoid GBM CSF samples were allocated to a discovery cohort, with the remainder of the samples (n=10/11) utilized for validation. Nearly identical overlap of the subarachnoid CSF proteome was seen between the discovery and validation cohorts (FDR=0.000; **Figure 1Aii**), with similar findings for discovery/validation analyses in the ventricular CSF proteome (FDR=0.000; **Figure 1Aiii**), confirming highly reproducible differences in these two locations.

**Figure 1.**
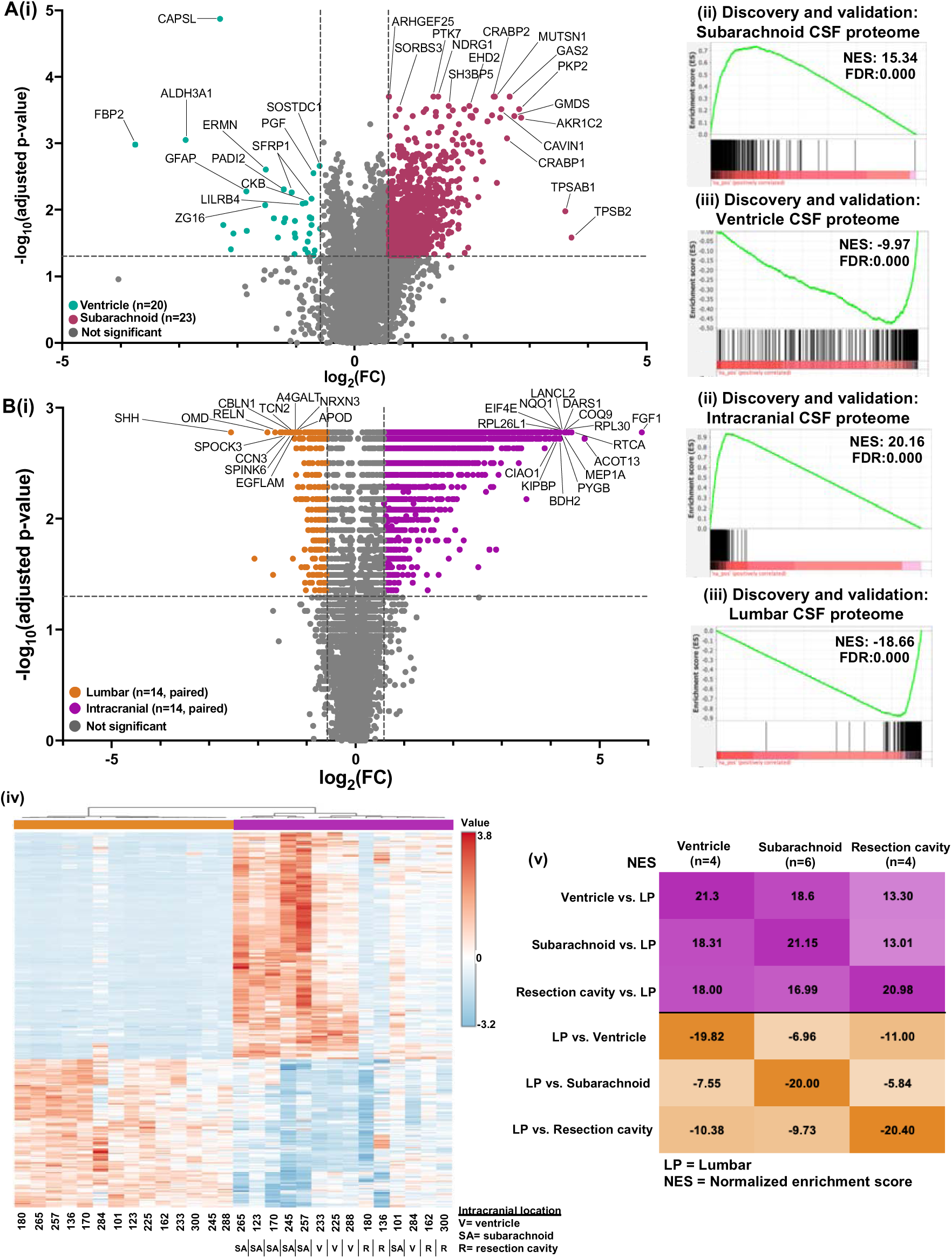
The CSF proteome differs by anatomical location. **1A.** (i) A volcano plot was generated by performing a series of Mann-Whitney U-tests with Benjamini-Hochberg correction on GBM CSF samples obtained from the ventricular system (n=20) versus the subarachnoid space (n=23). Cut-offs were FC≥1.5 and adjusted p-value≤0.05. (ii-iii) The ventricular and subarachnoid CSF samples were each split in half into discovery and validation cohorts (n=10 each in discovery and validation for ventricular; n=12 in discovery and n=11 in ventricular for the validation cohort), with subarachnoid versus ventricular CSF ranked fold-change lists generated for both cohorts. Enrichment analysis was performed to evaluate the enrichment of the discovery subarachnoid versus ventricular ranked fold-change list for the (ii) validation subarachnoid and (iii) validation ventricular CSF proteomes (top 350 proteins based on fold-change). **1B.** (i) A volcano plot was generated by performing a series of Wilcoxon-Signed rank tests with Benjamini-Hochberg correction on paired intracranial and lumbar CSF samples from patients with gliomas (n=14). (ii-iii) The lumbar and intracranial pairs were split in half into discovery and validation cohorts (n=7 pairs in each group), with paired intracranial versus lumbar ranked fold-change lists generated for both cohorts. Enrichment analysis was performed to evaluate the enrichment of the discovery intracranial versus lumbar ranked fold-change list for the (ii) validation intracranial and (ii) validation lumbar CSF proteomes. (iv) Hierarchical clustering was performed on the paired lumbar and intracranial CSF samples using the top 5% (350) proteins via t-test. (v) Ranked average fold-change lists were generated using the paired intracranial versus lumbar patient samples based on anatomical location (ventricle, subarachnoid, or resection cavity). Enrichment analysis was performed to evaluate the enrichment of each ranked list for proteomic libraries consisting of the top 5% (350 proteins) from the average paired intracranial versus LP lists. All FDR=0.000.

**Table 1.**
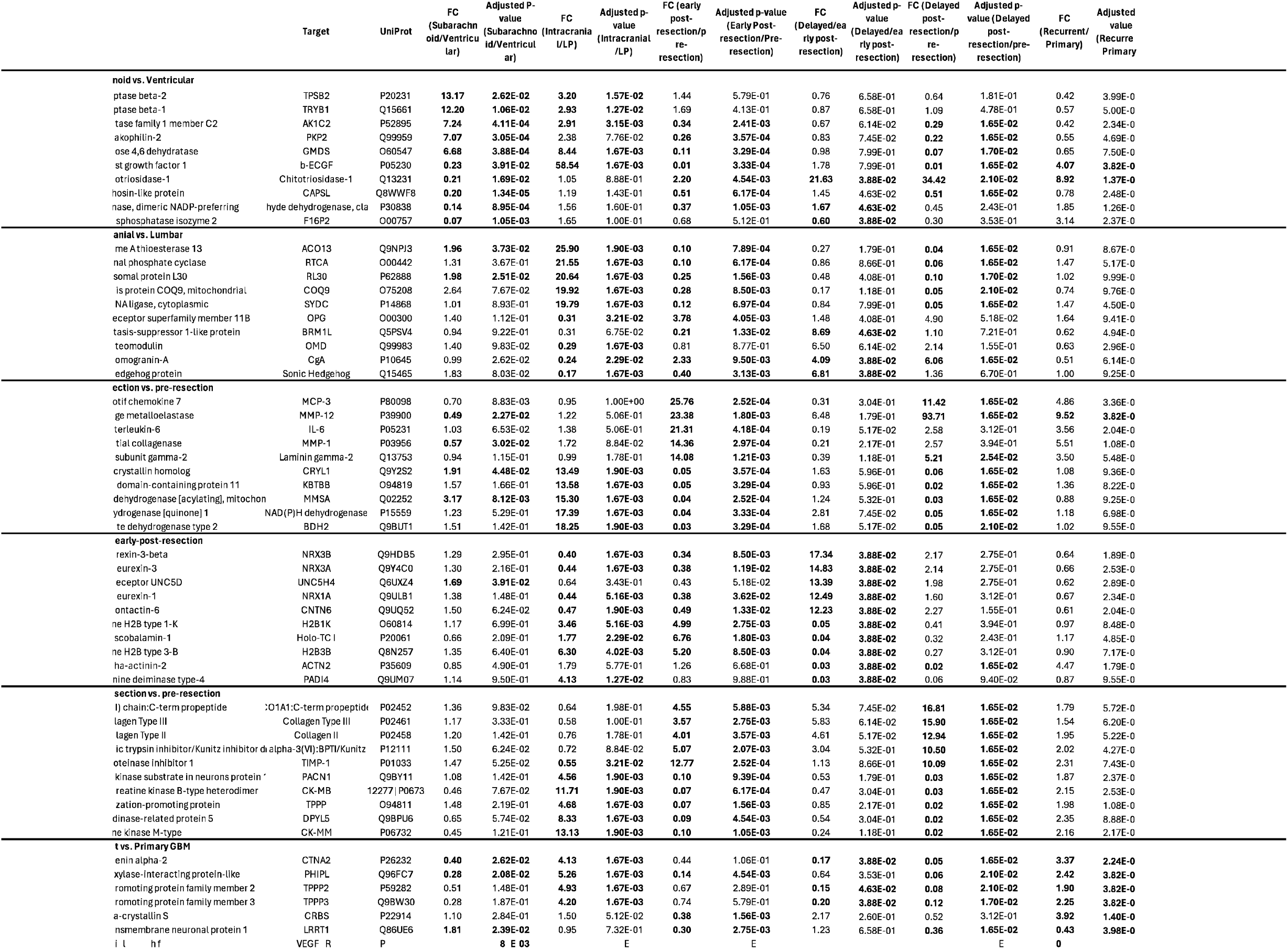
The Impact of Anatomical location and resection on the CSF proteome.

Having identified a proteomic impact from intracranial anatomic location, we hypothesized that a significant difference would exist between lumbar versus intracranial CSF. To address this question, we recruited patients to undergo intra-operative lumbar CSF sampling under anesthesia prior to surgery for intracranial tumor resection, when safe to do so. Among 14 paired intra-operatively acquired lumbar and intracranial CSF samples, 1,613 aptamers had an elevated signal in intracranial CSF, while 271 had an increased signal in lumbar CSF (FC≥1.5, adjusted p-value≤0.05; **Figure 1Bi**; **Table 1**; full list in **Supplementary Table 3**). Intracranial CSF was enriched for metabolism-related processes, while lumbar CSF was enriched for neuronal development pathways (**Supplementary Table 2**), consistent with increased abundance of SHH, a known craniocaudal patterning protein(*43*). Discovery and validation analysis (n=7 consecutive patients per cohort) demonstrated a highly reproducible location-associated CSF proteome via enrichment analysis and hierarchical clustering (FDR=0.000; **Figure 1Bii-iii)**. Hierarchical clustering revealed separated paired lumbar and intracranial samples based on anatomical location (**Figure 1Biv)**. We additionally asked whether the lumbar versus intracranial CSF signature was identifiable regardless of the site of intracranial CSF acquisition. Enrichment analyses of intracranial CSF samples based on location (ventricle, subarachnoid, or resection cavity) versus lumbar CSF revealed that the intracranial CSF proteomes were positively enriched for each other when compared to lumbar CSF (FDR=0.000 for most comparisons; **Figure 1Bv**), regardless of intracranial location. In sum, the intracranial CSF proteome across all sites of acquisition was distinct from the lumbar proteome.

### Longitudinal impacts of resection on the CSF proteome

During the patient’s disease course, the largest decrease in disease burden typically occurs via surgical resection. To determine the impact of surgical resection on the CSF proteome, paired pre- and post-resection intracranial CSF samples were acquired for twenty patients early in the patient’s post-operative course (median=10.5 days, range=1-35). Ranked fold-change lists were created to compare each patient’s pre-resection to their post-resection sample, revealing proteomic similarities in the pre-versus-post-resection signature across patients (**Figure 2Ai).** Notably, this consistent proteomic signature could be observed even in patients where post-resection CSF was obtained 3-5 weeks after resection as compared to within the first few days after surgery. To evaluate the reproducibility of these findings, the first and second 10 consecutive paired samples were utilized as discovery/validation cohorts, revealing strong reproducibility (FDR=0.000; **Figure 2Aii** and **Supplementary Figure 1A**). Given the impact of anatomical location (**Figure 1**), we also asked if the impact of resection could be observed independent of whether pre-resection CSF was obtained from the ventricular system (n=11) or subarachnoid space (n=7); two samples were excluded from analysis as the site of origin was unknown in one patient and another originated from a resection cavity. When compared to each patient’s post-resection CSF, ventricular and subarachnoid pre-resection CSF samples were enriched for one another, demonstrating a conserved impact of resection on the CSF proteome regardless of site of origin (FDR=0.000; **Supplementary Figure 1Bi-ii**). The same was also found when samples were divided based on primary (n=14) versus recurrence (n=6) status (FDR=0.000; **Supplementary Figure 1Ci-ii**).

**Figure 2.**
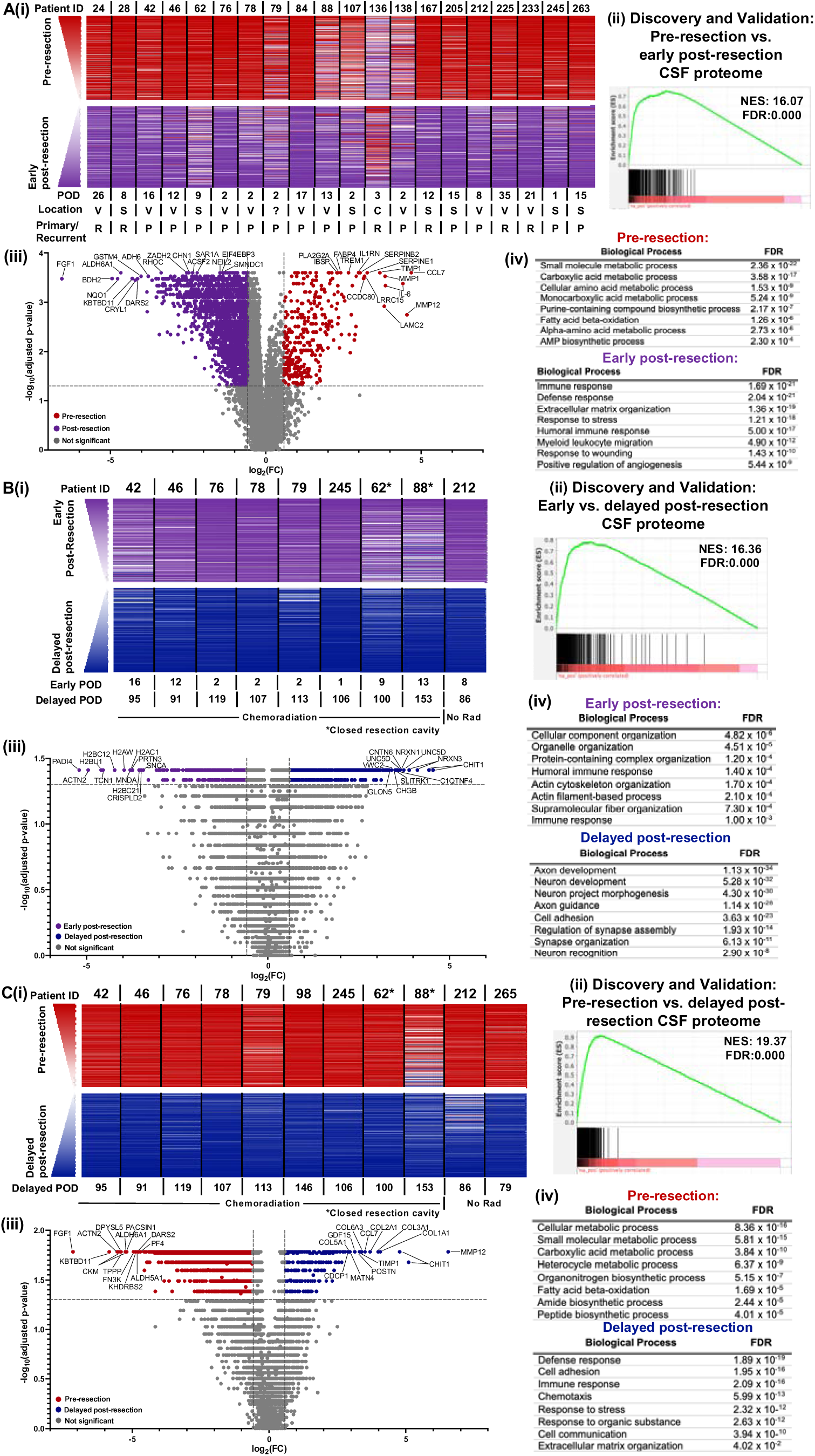
Glioma resection impacts the CSF proteome at early and delayed timepoints. **2A.** (i) Ranked fold-change lists were generated from each patient’s paired pre-versus-post-early resection (POD≤35) CSF samples (n=20). Ranked protein order is shown as a heatmap from 1 (higher pre-resection; red) to 7,011 (higher early post-resection; purple). The average rank across all 20 patients was used to list proteins. The top and bottom 200 proteins are shown. (ii) The 20 pre-versus-early post-resection CSF pairs were split in half into discovery and validation cohorts based on sequential order. Enrichment analysis of the discovery post- versus pre-early resection ranked fold-change list was performed for the validation pre-resection CSF proteome. (iii). A volcano plot was generated by performing a series of Wilcoxon Signed-Rank tests with Benjamini-Hochberg correction on the paired pre-versus- early post-resection CSF samples (n=20). Cut-offs were FC≥1.5 and adjusted p-value≤0.05. (iv) Functional protein network analysis was performed via STRING.db on the top 200 proteins significantly elevated prior to versus early after resection based on FC in 3Aiii. Gene ontology (GO) processes and their FDRs are reported. **2B**. Similar analyses were performed to 3A for the paired early-versus-delayed post-resection CSF samples (n=9 pairs). For (ii), the discovery cohort had 5 pairs of samples while the validation cohort had 4 pairs, based on sequential patient number. **2C**. Similar analyses were again performed to 3A-B, using the pre-resection versus delayed post-resection CSF samples (n=11 pairs). For (iii), the discovery cohort had 6 pairs of samples, while the validation cohort had 5 pairs, based on sequential patient order.

Wilcoxon rank-sum tests with Benjamini-Hochberg correction comparing the pre- versus post-resection samples demonstrated that the signal for 1,872 aptamers was significantly higher for CSF prior to resection, while the signal increased for 326 aptamers early after resection (adjusted p≤0.05; FC≥1.5); **Figure 2Aiii**; **Table 1**; full list in **Supplementary Table 4).** Functional protein network analysis of the top 200 of the 1,872 pre-resection aptamers revealed enriched gene ontology (GO) biological process terms for metabolism-related pathways, including small molecule metabolic processes (FDR=2.36×10^-22^) and fatty acid beta-oxidation (1.26×10^-6^) (**Figure 2Aiv)**. GO networks enriched in the 326 post-resection proteins included immune responses (1.69×10^-21^), extracellular matrix reorganization (1.36×10^-19^), and response to wounding (1.43×10^-10^), consistent with recent injury from surgical resection (**Figure 2Aiv)**.

We recently demonstrated that the extracellular glioma metabolome is enriched for plasma-associated metabolites, consistent with relative blood-brain barrier (BBB) disruption in contrast-enhancing regions(*44*). As resection should decrease the abundance of glioma-associated proteins, some of which could be due to BBB disruption, we asked whether resection could alter the relative enrichment of the CSF proteome for plasma-derived proteins. To do so, we compared the pre-versus-early post-resection proteome to the ranked list of plasma-derived proteins, generated from n=7 paired bloody versus clean CSF samples (**Supplementary Table 5)**. As expected, plasma-derived proteins were enriched for blood coagulation and immune system-related processes, as well as cellular organization processes (**Supplementary Table 2).** Early post-resection CSF is often more visibly contaminated than pre-resection CSF (**Supplementary Data**, annotations) due to post-surgical debris from recent tissue disruption. Nevertheless, plasma-associated aptamers were strongly enriched in glioma CSF at the time of maximal tumor burden when compared to early post-resection CSF (FDR=0.000; **Supplementary Figure 1Di)**, aligning with our recent findings in the glioma extracellular metabolome wherein blood-brain barrier portions of the tumor were enriched for plasma-derived metabolites(*44*). This finding suggests that certain proteins in the CSF glioma proteome may be impacted by plasma-derived proteins that cross the tumor-disrupted BBB.

Radiation with concurrent TMZ is standard-of-care after maximal safe resection for most patients with high-grade gliomas(*2*). We initially sought to characterize the impact of chemoradiation on the CSF proteome after resection in patients with primary gliomas. Evaluation of 8 patients’ pre-versus-post-chemoradiation CSF samples seemed to indicate a proteomic difference between samples obtained prior to versus after radiation. However, paired samples from one patient who did not receive chemoradiation revealed nearly identical proteomic changes during this timeframe (**Supplementary Figure 2A)**. We therefore hypothesized that the CSF proteomic changes occurring between these early (<35 day) versus late (>70 day) timepoints were more strongly attributable to the ongoing evolution of dynamic post-operative changes, than chemoradiation (**Figure 2Bi)**. Of note, evaluation of early and delayed timepoints was limited to patients with primary gliomas to minimize heterogeneity that could be induced from various times since prior resection and/or systemic treatments.

As with the pre- versus early post-resection analyses, the first five and second four patients were split into discovery and validation cohorts, respectively. Enrichment analysis demonstrated a reproducible impact of delayed post-resection changes on the CSF proteome as compared to the early post-resection timepoint (FDR=0.000; **Figure 2Bii; Supplementary Figure 3B)**. An elevated signal was detected for 268 aptamers in the early post-resection setting while 443 aptamers had an increased signal at the delayed timepoint (**Figure 2Biii**; **Table 1**; full list in **Supplementary Table 6).** Pathways related to cellular component organization (FDR=4.82×10^-6^), organelle organization (4.51×10^-5^), and immune responses (1.40×10^-4^) were more abundant in the early post-resection setting (**Figure 2Biv)**. At the delayed timepoint, GO processes were enriched for neuronal/synaptic related pathways, including axon development (1.13×10^-34^), regulation of synapse assembly (1.93×10^-14^), and neuron recognition (2.90×10^-8^) (**Figure 2Biv)**. Moreover, enrichment for plasma-derived proteins decreased by the delayed post-resection timepoint (**Supplementary Figure 2C**). Overall, these data suggested an increased abundance of neuronal-associated proteins as post-resection changes evolve over time.

Having compared pre-resection to early post-resection CSF, we then asked how the former would compare to the delayed post-resection timepoint. Comparing the 11 paired pre-resection versus delayed post-resection CSF samples resulted in similar findings (**Figure 2C)** from the pre-resection versus early post-resection comparison (**Figure 2A; Supplementary Table 7)**. These findings included a highly reproducible signature across discovery/validation cohorts (FDR=0.000; **Figure 2Cii; Supplementary Figure 3A)** and GO processes enriched for metabolism-associated pathways prior to resection and for immune/stress responses at the delayed post-resection point (**Figure 2Ciii-iv**). Likewise, pre-resection CSF was more strongly enriched for plasma-derived proteins than the delayed post-resection timepoint (FDR=0.000; **Supplementary Figure 3B**). In sum, the impacts of resection could be identified in the CSF proteome at both early and delayed timepoints, although neuronal-associated proteins became more abundant with more time since resection.

### Evolution of post-resection CSF proteomic changes

After comparing two of the three timepoints to one another, we then sought to integrate these findings to evaluate the dynamic evolution of the CSF proteome. To do so, we identified proteins that were more abundant at one timepoint as compared to both of the other timepoints (**Figure 3Ai; Supplementary Table 8**). These analyses identified a subset of proteins that (i) were elevated pre-resection and remained low post-resection (row 1 in **Figure 3Ai**; example: fibroblast growth factor 1, **Figure 3Bi**), (iii) increased transiently in the early post-resection setting (row 2; example: histone H2B Type 3-B, H2BU1, **Figure 3Bii**), (iii) increased early post-resection and remained elevated by the delayed post-resection timepoint, and (iv) increased most significantly in a delayed fashion post-resection (row 3; example, growth differentiation factor-15 (GDF-15), **Figure 3Biii**) (**Supplementary Table 8**). Additionally, evaluation of the top 10 plasma-associated proteins demonstrated decreased abundance over time (**Figure 3Aii**), consistent with decreasing enrichment for the plasma-associated signature from pre-resection to delayed post-resection CSF. Distinct differences were noted in the functional protein network enrichments at each timepoint. Namely, pre-resection proteins were enriched for metabolism-related processes and the early and delayed post-resection for stimuli response/immune processes pathways (**Supplementary Table 2**).

**Figure 3.**
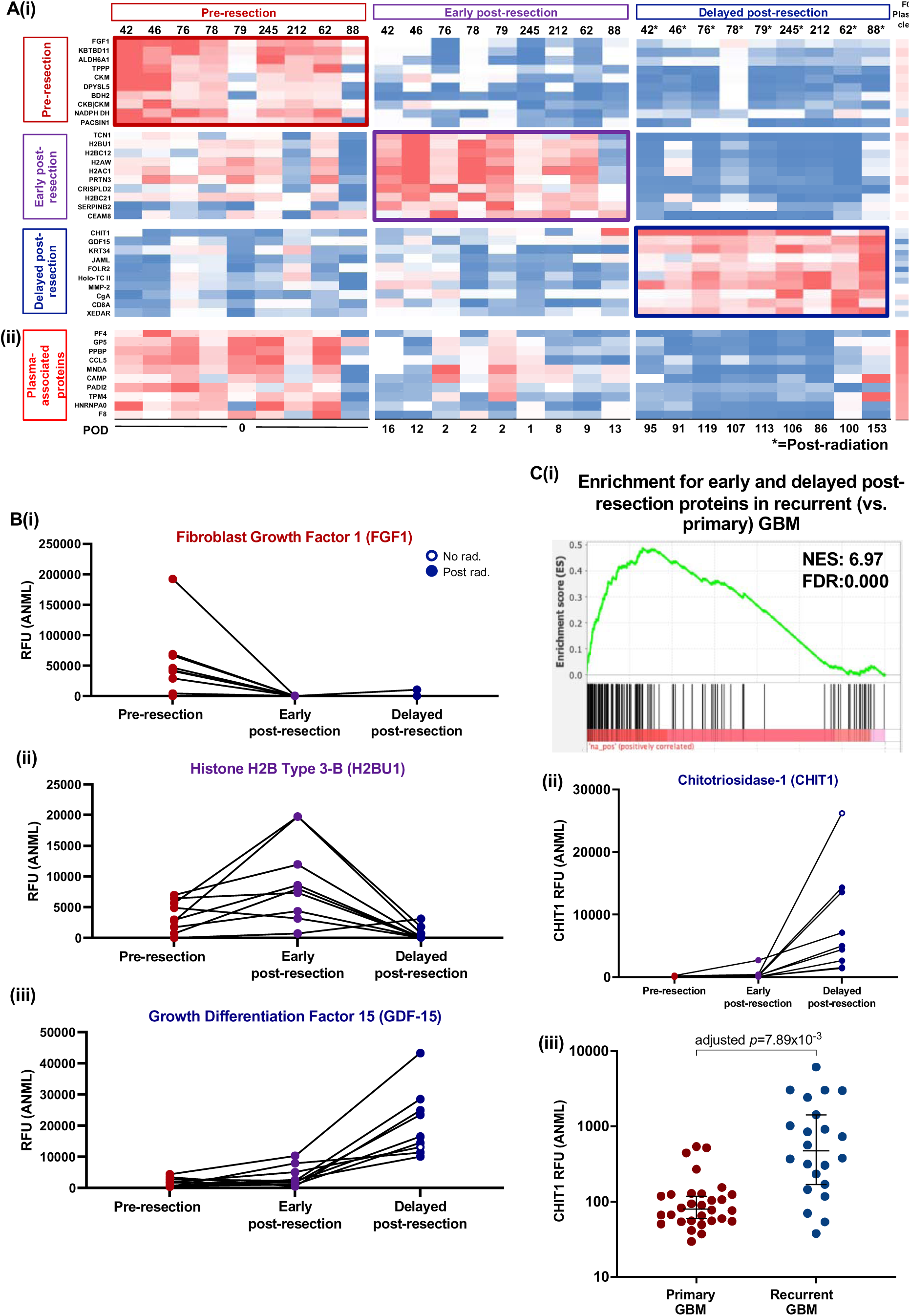
The impact of resection evolves over time and is detectable at recurrence. **3A**. (i) The abundance of the top 10 proteins unique to each timepoint are shown for 9 patients with gliomas who had CSF samples acquired intra-operatively, early post-resection (POD≤35), and delayed post-resection (POD≥70). Proteins unique to each timepoint were found by identifying the proteins that were significantly elevated (FC≥1.5, adjusted p≤0.05) at that timepoint when compared to both of the other timepoints. The fold-change in bloody versus clean CSF samples (n=7 pairs) is also shown. (ii) The abundance of the top 10 plasma-associated proteins are shown for these patients’ samples. **3B**. The abundance at each timepoint for each patient is shown for (i) Fibroblast growth factor 1 (FGF1), (ii) Histone 2B Type 3-B (H2BU1), and (iii) growth differentiation factor 15 (GDF-15), which were in the top 10 proteins at the pre-resection, early post-resection, and delayed post-resection timepoints, respectively. **3C**. (i) Proteins were identified that were significantly elevated in both the early and delayed post-resection timepoints when each were compared to the pre-resection timepoint (FC≥1.5 and adjusted p≤0.05). 150 proteins overlapped between these two comparisons. Enrichment analysis was performed to evaluate the enrichment of the recurrent versus primary GBM proteome ranked list for these 150 proteins that were elevated at both early and delayed post-resection timepoints as compared to pre-resection. (ii-iii) Chitotriosidase 1 (CHIT1) was evaluated in the pre-resection, early post-resection, and delayed post-resection timepoints, as well as in primary (n=31) versus recurrent (n=21) GBM samples acquired intra-operatively.

Moreover, we also investigated whether proteins that increased over time after resection would remain elevated at the time of resection for recurrent glioma. To do so, we compared the unpaired proteome of primary (n=31) versus recurrent (n=21) GBM, which identified 18 aptamers with significantly increased signal in the recurrent subgroup (FC≥1.5, adjusted p-value≤0.05) (**Table 1**; full list in **Supplementary Table 9**). Enrichment analysis revealed that proteins elevated at the delayed post-resection timepoints (including its overlap with the early post-resection proteins; n=150 aptamers) were enriched in the recurrent GBM CSF proteome when compared to primary GBMs (FDR=0.000; **Figure 3Ci**). Indeed, chitotriosidase-1 (CHIT1) was the protein that most significantly increased after resection (**Figure 3Ci)** and was the second most abundant protein in recurrent GBM when compared to primary tumors (**Figure 3Cii**). Although there was a low number of significantly differentially enriched proteins by recurrence after p-value adjustment for multiple hypothesis testing, the significant proteomic enrichment for the post-resection signature suggests that its inflammatory components may still be detectable by the time of recurrence. In sum, the CSF proteome of gliomas dynamically evolves over time with resection.

### The dynamic CSF proteome during therapy

Beyond imaging, few methods exist for monitoring glioma disease burden. We hypothesized that proteins decreasing with glioma resection could plausibly serve as an initial protein set from which to identify candidate monitoring biomarkers. To assess the pilot feasibility of this approach, we evaluated the relative abundance of the top 25% of proteins decreasing with resection (**Figure 1A)** in three patients with grade 4 gliomas who exemplified divergent disease courses over time. Patient 24, a female in her 50s, progressed through multiple treatments at recurrence after resection of a recurrent GBM (**Figure 4A)**. Patient 46, a female in her 40s, had stable disease burden after resection and chemoradiation of an IDH-mutant grade 4 astrocytoma (**Figure 4B)**. Patient 98, a male in his 40s, demonstrated pseudoprogression following resection and chemoradiation of an IDH-mutant grade 4 astrocytoma (**Figure 4C)**.

**Figure 4.**
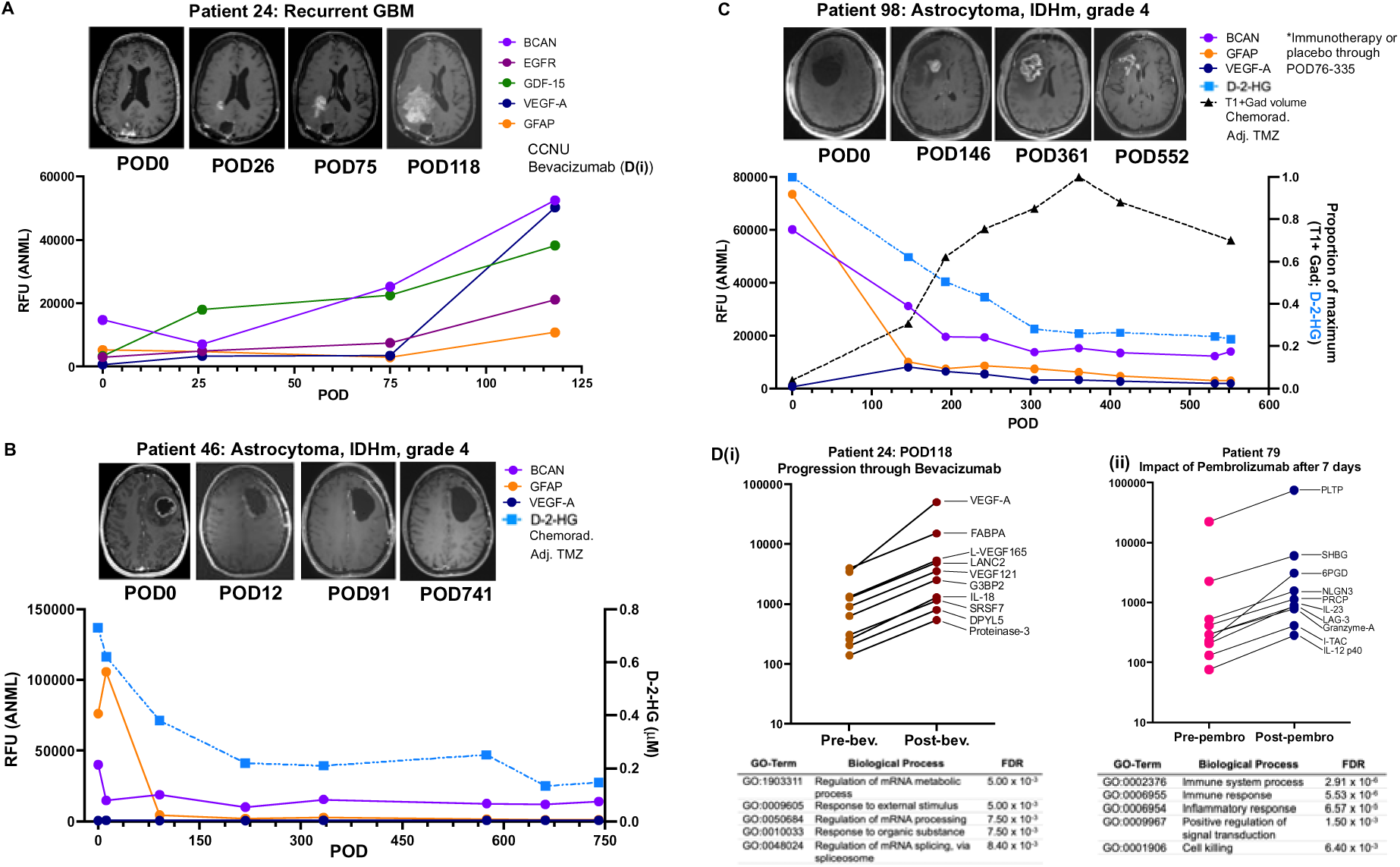
Intracranial CSF can be acquired longitudinally for evaluation of candidate monitoring biomarkers and pharmacodynamic impact of therapies. **4A**. BCAN, epidermal growth factor receptor (EGFR), growth differentiation factor-15 (GDF-15), VEGFA-A, and GFAP were evaluated in intracranial CSF from patient 24, who had a recurrent GBM with known EGFR amplification. Green box = days on CCNU; maroon box = days on bevacizumab. **4B.** The normalized RFUs of brevican (BCAN), glial fibrillary acidic protein (GFAP), and vascular endothelial growth factor-A (VEGF-A) were evaluated over time in longitudinal intracranial CSF obtained from a patient with an astrocytoma, IDH-mutant, grade 4 via an Ommaya reservoir. D-2-hydroxyglutarate (D-2-HG) was also evaluated at each timepoint. Red box = days during which patient underwent chemoradiation; yellow box = days on adjuvant temozolomide (adj. TMZ). **4C**. BCAN, GFAP, and VEGF-A were evaluated in longitudinal intracranial CSF from a patient with an astrocytoma, IDH-mutant, grade 4, as well as D-2-HG. Additionally, the T1-post gadolinium positive (T1+Gad) volume was calculated from MRIs obtained at each timepoint. Red box = days during which patient underwent chemoradiation; yellow box = days on adjuvant temozolomide (adj. TMZ). **4D**. The top 10 proteins based on fold-change are shown for post-versus-pre-treatment with (i) bevacizumab in patient 24 from 4A, as well as (ii) post-versus-pre-pembrolizumab in patient 79, who had a GBM with a hypermutated phenotype.

As an initial strategy, we reasoned that a monitoring biomarker should increase with true disease progression. Of these top 25% proteins, BCAN emerged as the strongest candidate in patient 24 based on its fold-change between progression timepoints, increasing by 3.58x and 2.08x during progression through lomustine and bevacizumab, respectively (**Figure 4A)**. Most of the other proteins elevated prior to resection did not increase reliably with disease progression at each timepoint (**Supplementary Figure 4Ai).** Of note, VEGF-A had the greatest increase in signal of all aptamers elevated with progression through bevacizumab (14.45x), consistent with its relevance to blood-brain barrier disruption in HGGs(*45, 46*) in the setting of failed radiographic response to this anti-VEGF-A antibody (**Figure 4A)**. GFAP was also evaluated as a previously proposed biomarker of GBM(*47–49*) and one of the top proteins that increased with progression through bevacizumab (3.59x). However, unlike BCAN, both VEGF-A and GFAP did not increase with progression through lomustine (1.03x and 0.63x, respectively). Epidermal growth factor receptor (EGFR) and growth-differentiation factor 15 (GDF-15), an indicator of p53 activity(*50*), were also evaluated for patient 24, since her GBM was EGFR-amplified and P53-wild type. EGFR and GDF-15 changed nominally during progression through lomustine, but increased by 2.83x and 1.70x, respectively, with progression through bevacizumab.

We then turned to evaluating BCAN, GFAP, and VEGF-A in patient 46, who had a radiographically stable IDH-mutant grade 4 astrocytoma after resection and chemoradiation. EGFR and GDF-15 were not evaluated due to their lack of relevance to her tumor genomics. In patient 46, the BCAN signal decreased by 0.61x by the end of adjuvant temozolomide at POD334 (**Figure 4B),** consistent with decreasing D-2-hydroxyglutarate (D-2-HG), an oncometabolite of IDH-mutant gliomas. GFAP increased by 1.39x with resection and decreased by 0.96x from pre-resection after adjuvant temozolomide, while VEGF-A remained low since baseline. All proteins, including those that decreased with resection (**Supplementary Figure 4Aii),** remained relatively unchanged thereafter, correlating with her stable imaging.

Finally, in patient 98 whose disease course was characterized by pseudoprogression, signals for BCAN and GFAP decreased to 0.23x and 0.10x of baseline, respectively, by POD305 after standard-of-care chemoradiation with adjuvant temozolomide (**Figure 4C),** correlating with decreasing D-2-HG. In contrast, VEGF-A increased by 12.67x between the post-chemoradiation and intra-operative baseline sample, as did a number of other proteins elevated prior to resection that either then stabilized or began decreasing (**Supplementary Figure 4Aiii)**. Indeed, while BCAN, GFAP, and VEGF decreased with each subsequent CSF sample, the volume of T1-gadolinium positive tumor increased over time, until it began to decrease by POD552, likely indicative of pseudoprogression.

Additionally, we also sought to test the preliminary feasibility of detecting pharmacodynamic signals in CSF during systemic treatment. Protein network enrichment analysis of patient 24’s CSF proteome during progression through CCNU between POD26 and 75 revealed similar neuronal-related enrichment pathways to the early versus delayed post-resection signature, consistent with a signature of resolving post-surgical changes between those timepoints and suggesting minimal pharmacodynamic impact of CCNU **(Supplementary Figure 4B-4Ci)**. In contrast, numerous aptamers increased during progression through bevacizumab between POD75 and 118, including VEGF-A (14.45x), and its isoforms, VEGF165 and VEGF121 (**Figure 4Di)**. GO terms for progression through bevacizumab included mRNA metabolic processes (5.00×10^-3^), response to organic substance (7.50×10^-3^), and regulation of mRNA splicing (8.40×10^-3^). Increases in these proteins between POD75 and 118 contrasted their decrease in other patients over time after surgery (**Supplementary Figure 4Cii**), suggesting a distinct PD impact of bevacizumab in CSF.

In another patient (Patient 79), CSF samples were obtained immediately prior to and 7 days after receiving pembrolizumab for a primary GBM with a hypermutant phenotype (POD28-36). Observations included increased abundance of diverse interleukins, as well as lymphocyte activation gene-3 (LAG-3), which is associated with T cell activation and serves as an alternate immune checkpoint to PD1(*51*), as blocked by pembrolizumab (**Figure 4Dii).** STRING analysis demonstrated increased immune system processes (FDR=2.91×10^-6^) and cell killing pathways (6.40×10^-3^). Again, the increases in these proteins differed from their longitudinal time course in other patients (**Supplementary Figure 4Ciii**), potentially indicative of a pharmacodynamic impact of pembrolizumab.

Overall, these data suggest that the CSF proteome may reflect biological changes occurring during disease evolution and therapy. Further work will be needed to evaluate the reproducibility of such changes across patients and to determine the performance of other alternatives, such as individualized protein signatures based on each patient’s baseline proteome, for disease monitoring.

## DISCUSSION

Toward our goal of understanding variables impacting the longitudinal glioma CSF proteome, namely anatomical location and resection, we sampled intracranial or lumbar CSF at resection, in clinic, and/or longitudinally via CSF access devices in 74 patients comprising a total of 146 samples, 71 of whom had grade 2-4 astrocytomas or grade 2-3 oligodendrogliomas. Our results demonstrate (1) a substantial impact of anatomical location on the glioma CSF proteome, namely between subarachnoid versus ventricular CSF that may have implications for longitudinal CSF acquisition wherein post-operative samples are often obtained from CSF in continuity with the ventricle, (2) resection-induced changes in the intracranial glioma CSF proteome that evolve over time, (3) enrichment for plasma-associated proteins prior to resection that decreases temporally after resection, and (4) feasibility of longitudinal CSF acquisition for analysis of preliminary pharmacodynamic changes in the intracranial CSF proteome. Although many questions remain unanswered, our available data to date suggest that the intracranial glioma CSF proteome provides an abundant and evolving source of candidate biomarkers to longitudinally discern the impact of resection and systemic treatments. All data are provided as a resource for the neuro-oncology community to foster efforts to communally answer these remaining questions and identify glioma biology via CSF proteomics.

While tissue is rarely accessible after therapy for treatment response assessment(*52*), CSF can be sampled at multiple timepoints. We identify a conserved impact of resection on the glioma CSF proteome that is consistent with a tissue injury-induced inflammatory milieu(*53, 54*), and which persists in the months after surgery (**Figure 2-3).** Increased enrichment for neuronal-associated pathways between the early (<1 month) to delayed post-resection timepoint (2-5 months) suggest the ability to recover the brain CSF proteome after resection. This observation could be due to decreased tumor burden with resection and chemoradiation. Additionally, decreasing enrichment for plasma-associated proteins over time may contribute to this finding(*55*), as non-plasma-associated CSF proteins in our paired bloody-versus-clean samples shared similar functional pathways related to cell adhesion and system/neuron development, albeit to less significant levels (**Supplementary Table 2)**. Additional longitudinal CSF samples from patients with low-grade gliomas, as with patient 212 (**Figure 2**), not undergoing any therapy after resection will be utilized to isolate evolving post-resection changes from those induced by chemoradiation, other therapies, and disease recurrence. Although far more uncommon, non-treated, post-resection samples from patients with HGGs would also be of use since the difference in the tumor microenvironment between LGGs and HGGs could plausibly impact these evolving changes.

Moreover, the pre-resection CSF proteome was highly enriched for plasma-derived proteins which decreased overtime after resection. While decreased plasma-derived enrichment between early and delayed post-resection samples may be attributable to clearing of surgical products, there was visibly increased contamination early after resection in longitudinal CSF samples as compared to baseline pre-resection samples, where plasma-derived protein enrichment was the highest. A prior CSF proteomic study found that abundance of albumin, a plasma-derived protein, correlated with the number of differentially abundant CSF proteins(*33*). Our prior findings via microdialysis suggest that the tumor microenvironment is more enriched for plasma-derived metabolites in the BBB disrupted as compared to BBB intact portions of the tumor(*44*). As such, it is possible that tumor-associated blood-brain barrier disruption may contribute to a portion of the glioma proteome. Indeed, clean intracranial CSF samples (n=6) were significantly more enriched for plasma-associated proteins than their paired lumbar counterparts (**Supplementary Figure 1E**), consistent with the tumor as a source of plasma-associated proteins. The impact of BBB disruption could in part contribute to the large number of proteins that were more abundant prior to resection due to tumor-induced BBB disruption, as compared to immediately post-resection (1,872 versus 326 proteins, respectively; **Figure 2A**). Whether plasma-derived protein enrichment has any impact on glioma growth and proliferation remains to be determined. However, serum is a routine component of cell culture media(*56*). Future studies may seek to evaluate whether blood-derived mitogens crossing a disrupted BBB could be as protumorigenic for the human glioma, *in situ*, as it is for *in vitro* cell cultures.

Our findings suggest that the anatomical site of CSF origin, including lumbar or subarachnoid versus ventricular CSF intracranially, profoundly impacts the CSF proteome. Previous work demonstrated a significant difference in the composition of paired intracranial versus lumbar CSF in patients with NPH(*37*). To our knowledge, similar findings have not previously been demonstrated for patients with gliomas. Cranial CSF should presumably afford a more abundant source of glioma-associated proteins due to tumor proximity(*11, 57*). Indeed, our data in paired lumbar and intracranial glioma CSF samples demonstrate more differentially abundant proteins in intracranial CSF than lumbar CSF (**Figure 1B),** although this proximity may also increase the abundance of brain-associated proteins. Of note, of these 1,613 aptamers with higher signal in intracranial than lumbar CSF, 1,143 decreased with resection (70.9%; **Supplementary Table 10**), some of which could plausibly be associated with gliomas, including its microenvironment and BBB disruption.

Differences in the subarachnoid versus ventricular CSF should also be considered for longitudinal evaluation of samples due to changes in tumor contact, diffusion distance into CSF, and continuity with the ventricular system that could impact protein abundance. However, this is likely to depend on the protein under evaluation. For example, of 1,872 aptamers with a higher signal prior to resection as compared to early post-resection, 696 (37.2.%) were also associated with the subarachnoid GBM CSF, but 1,168 (62.4%) were not significantly different between ventricular versus subarachnoid GBM CSF (**Supplementary Table 10**). Of note, the ventricular system is frequently reached intra-operatively later than the subarachnoid CSF, after surgical tissue disruption has occurred that could increase sample bloodiness. As such, we originally anticipated that ventricular CSF would be more enriched for plasma-associated proteins than subarachnoid CSF. However, we noted greater enrichment of subarachnoid GBM CSF for plasma-associated proteins when compared to ventricular CSF (**Supplementary Figure 1F)**. Subarachnoid CSF is more often sampled when there is greater tumor burden present (often prior to resection). As many proteins elevated prior to glioma resection are plasma-associated, a potential hypothesis may be that its enrichment for plasma-associated proteins originates from greater tumor burden present during sampling, although this remains to be proven.

Despite such need for careful consideration of potential co-variates, our data suggest the feasibility of discerning unique biological impacts of tumor-targeted therapy within a patient’s CSF by serially sampling same location prior to and after therapy in individual patients (**Figure 4D)**. Pembrolizumab increased the abundance of numerous cytokines/chemokines, as expected from an immunotherapy--especially in a hypermutant tumor(*58*). Interestingly, LAG-3, an immune checkpoint inhibitor, was more abundant in CSF after blockade of the PD-L1 checkpoint inhibitor pembrolizumab. Combined blockade of PD-L1 and LAG-3 improved survival in preclinical models and is currently under clinical trial evaluation for GBM, illustrating the potential of identifying emergent mechanisms of resistance via CSF analysis on a patient-by-patient basis(*59*). Of note, while LAG-3 did not increase with resection when compared to pre-resection, it did increase by the delayed post-resection timepoint as compared to early pre-resection. As such, the temporal changes occurring in CSF after resection, independent of these targeted therapies, should be considered in parallel when evaluating PD impact. In contrast, progression through CCNU and bevacizumab in one patient yielded two distinct CSF proteomic signatures, perhaps illustrating the dynamic nature of tumor responses to therapeutic stress. The >20x upregulation of VEGF-A as the top upregulated protein following bevacizumab may illustrate the tumor ecosystem’s capacity to resist therapy with remarkable precision. Again, parallel timepoints in patients with recurrent disease not undergoing treatment with CCNU or bevacizumab are likely needed to definitively attribute these changes to the therapies. Nevertheless, these data provide early signals on the potential of CSF for discerning PD impacts toward empirically guided individualized therapy. In sum, the data provided in this manuscript illustrate the potential utility of leveraging the dynamic CSF proteome longitudinally in our collective efforts against glioma.

## MATERIALS AND METHODS

### Patient recruitment, informed consent, and IRB approval

Cerebrospinal fluid (CSF) samples were collected with informed consent via one or more Institutional Review Board-approved protocols. Samples collected intra-operatively from the surgical field were banked under our neuro-oncology biorepository protocol. Any patient undergoing resection was eligible for enrollment into the biorepository protocol. Additionally, longitudinal CSF sampling was enabled in most cases by an Ommaya reservoir implanted under a research protocol for brain tumor biomarker access at the time of tumor resection (NCT04692337). For this initial study, patients with suspected or known low-grade or high-grade gliomas were recruited. Moreover, some samples were accessed via a clinically-indicated ventriculoperitoneal shunt or post-operative external ventricular drain under our CSF biomarkers study (NCT04692324). The CSF biomarkers study was also utilized to obtain lumbar CSF from lumbar punctures, either intra-operatively after anesthesia induction or in clinic.

### Cerebrospinal fluid collection and processing

Intra-operatively obtained samples were most often acquired from an exposed sulcus after dural opening, a prior resection cavity, and/or once the ventricle was reached, when applicable. Efforts were taken to minimize blood contamination of each sample. For patients with an Ommaya reservoir, the catheter was placed in the resection cavity at the completion of the case—most of which contacted the ventricle, except where annotated in the figures. Most Ommayas utilized had a 1.5 cm side with a flat bottom and side inlet connected to an antibiotic-impregnated ventricular catheter (Natus ® NT8501214; Codman ® Hakim Bacisteal Catheter, NS5048). Ommayas were placed such that would be minimally visible while ensuring the catheter did not cross the incision line. For all CSF samples, CSF was centrifuged at 400xG for 10 minutes at 4°C and aliquoted for storage at −80°C, with collection and processing time noted for each sample. Clinical information, including pathology, age, sex, tumor location, and other variables, was documented for each sample (“Annotations”, **Supplementary Data**).

### Aptamer-based proteomics

De-identified CSF samples were sent for aptamer-based proteomics analysis on the Somalogic SomaScan® platform(*40*). This method utilizes protein-captured Slow Off-rate Modified Aptamers (SOMAmers ®) to sensitively and reproducibly (<5% coefficient of variance) detect over 7,000 proteins in liquid specimens. Some of these single-stranded DNA-based SOMAmer reagents have been chemically modified and incorporated into nucleotide libraries for selection and amplification via Systematic Evolution of Ligands by EXponential enrichment (SELEX)(*41*). Although originally utilized for human plasma, serum, and urine, this technology is now being deployed in CSF(*60*).

Details of this method have been more extensively detailed in prior publications(*40*). Briefly, 20 uL of CSF are diluted and incubated with SOMAmer reagent mixes including streptavidin (SA)-coated beads. After the beads are washed, an NHS-biotin agent is utilized to tag the proteins specifically bound by SOMAmer reagents. Ultraviolet light then cleaves the linker within the SOMAmer reagent to release the SOMAmer complex and unbound reagents into an anionic solution, causing any nonspecific interaction to dissociate and preventing reformation. Beads are then used to separate the photocleavage eluate prior to incubation with a second set of SA-coated beads which bind biotin-labeled proteins and those complexed with SOMAmers. Another washing step is utilized to remove non-specifically bound reagents. Finally, protein-bound SOMAmer reagents are released via denaturation to quantify the reagents via hybridization to custom DNA microarrays based on a cyanine-3 signal in the reagent.

### Data standardization and processing

The 96-well plate utilized by Somalogic includes control, quality (pooled), calibrator (pooled), and buffer (no protein) samples. Raw data obtained from hybridization undergo hybridization normalization. Hybridization normalization uses twelve control SOMAmer reagents that are added to the eluate prior to scanning the microarray, without exposure to proteins. Median signal normalization is then performed using pooled calibrator replicates to normalize for within-run technical variation. Then, for each SOMAMer® reagent, a scaling factor is calculated and performed, including 1) the plate scale (median ratio; to adjust for differences in signal intensity between runs) and 2) the calibration scale (recalculated set of scale factor based on calibrators; to adjust for SOMAmer reagent assay differences between runs). A scaling factor is then applied to the microarray based on a scaling factor from the control signals. Adaptive Normalization by Maximum Likelihood (ANML) normalization is then performed. Each SOMAmer reagent has a scale factor within a dilution bin (S1, S2, S3) that is calculated based on sample values within two population standard deviations of the normal reference. This process is then repeated until convergence to increase the likelihood that the RFU for a sample is from the reference distribution.

Samples were sent across multiple batches. To evaluate for potential batch effects, a small subset of samples was sent across multiple batches. Data were provided by Somalogic for each step in their analysis pipeline. As another normalization technique, we had also performed 1) Trimmed Mean of M-values normalization(*61*) (based on top and bottom 5%) and 2) median normalization based on the median value within a sample on Somalogic’s hybridization normalized data. To determine which data normalization strategy to utilize for our analyses, we evaluated the correlation of our duplicate samples across batches to one another. Overall, ANML normalized data had the strongest correlation between batches for repeat samples (R^2^= 0.94-0.97, 5 samples) and was utilized for all analyses hereon. Proteins flagged in any batch were removed from downstream analyses, resulting in 7,011 analyzable aptamers compared to a starting point of 7,596 aptamers.

### Enrichment analyses

To perform enrichment analyses, the ANML value for each aptamer was compared across samples. In order to compare paired samples for an individual patient, aptamers were ranked based on fold-change (sample 1 / sample 2) resulting in a ranked list for each comparison (e,g pre- versus post-resection). For group comparisons, the average ANML value for each group was calculated and the fold change of each protein between groups determined to generate the ranked list. For discovery and validation analyses, samples were split into half, with the first chronological half (based on patient number) in the discovery cohort and the second half in the validation cohort. Discovery and validation were also performed based on primary versus recurrent tumor status and subarachnoid versus ventricular pre-resection CSF, where applicable in the manuscript. Fold-change ranked lists were created for each discovery and validation cohort. If a patient had multiple intra-operative CSF samples obtained, or if their CSF was run across multiple batches, the first CSF sample obtained was utilized under the most realistic assumption that only one sample could be obtained in most surgeries. The only exception was if the first sample was part of known bloody and clean pair of samples, in which case the less contaminated sample was utilized.

To determine the statistical significance of the similarity between ranked lists, Gene Set Enrichment Analysis® was repurposed for proteomics. Libraries for each comparison were generated using the top 5% of proteins in the comparison based on the fold-change (350 proteins). The exception was the evaluation of early and delayed post-resection proteins in the recurrent versus primary GBM proteome, that were identified as described in *Volcano plots, heatmaps, and hierarchical clustering* below. The Ranked lists (.rnk) were then utilized to determine where the proteins in each library fell on the ranked list. Ranked lists were based on fold-changes across comparisons of interest. The pre-rank function was utilized with classic enrichment statistic, mean-divided normalization, and 1000 permutations. Positive enrichment indicated that the library’s proteins fell toward the top of the list, suggesting an overlap in their upregulated proteomes. Negative enrichment indicated that the ranked proteome was similar to the opposite end of the ranked list.

### Volcano plots, heatmaps, and hierarchical clustering

Volcano plots were generated based on results of non-parametric t-tests performed to compare groups for each aptamer. First, normality of the overall data set was tested via D’Agostino-Pearson test. As the data were not normally distributed for every aptamer, non-parametric testing was performed for all aptamers, including Wilcoxon Signed-rank tests for paired data (pre-versus post-early or delayed resection; early versus delayed post-resection; lumbar-versus-intracranial) or the Mann-Whitney U tests for unpaired data (ventricular versus subarachnoid GBM; primary versus recurrent GBM). Given the number of aptamers on the panel, multiple hypothesis correction was performed using the Benjamini-Hochberg method.

For the heatmap in Figure 3A, the top 10 proteins were identified for each row from the volcano plots generated in Figure 2 at FC≥1.5 and adjusted p-value≤0.05. To identify proteins unique to one timepoint, we evaluated the proteins that were elevated in that timepoint when it was compared to both of the other timepoints. For example, to identify pre-resection specific proteins, we identified the significant proteins overlapping between the pre-resection versus early-post-resection and pre-resection versus delayed post-resection volcano plots. To identify proteins shared between two timepoints, but not the other, we evaluated the overlap between those two timepoints as compared to the third timepoint. For examples, to identify proteins shared between pre-resection and early post-resection samples, but not delayed post-resection CSF, we found the significant proteins overlapping between pre-resection versus delayed-post resection and early post-resection versus delayed-post-resection comparisons. The top 10 plasma-associated proteins based on the plasma-derived versus clean FC were plotted over time in each longitudinal samples.

STRING.db was utilized to perform functional protein network enrichment analyses and identify Gene Ontology (GO) processes by inputting up to the top 200 proteins for each protein list based on fold-change, or in the case of Figure 3/Supplementary Table 8, the proteins that were identified for each timepoint. MetaboAnalyst 6.0 was utilized to perform hierarchical clustering (Euclidean distance measure, with Ward’s method for clustering). ANOVA or t-tests were utilized for hierarchical clustering. GraphPad PRISM 10.0.1 was utilized to generate graphs.

## List of Supplementary Materials

Supplementary Figures S1: Enrichment analyses with paired pre-versus-early post-resection CSF samples from patients with gliomas.

Supplementary Figure S2: Enrichment analyses in 9 paired early and delayed post-resection CSF samples from patients with gliomas.

Supplementary Figure S3: Enrichment analyses in the 11 paired pre-resection versus delayed post-resection CSF samples from patients with gliomas.

Supplementary Figure S4: Evaluation of candidate monitoring and pharmacodynamic proteins in longitudinal CSF samples.

Supplementary Tables 1-10

Data File 1: all raw data

## Data Availability

All data produced in the present study are available upon reasonable request to the authors and will be published with the manuscript.

## ACKNOWLEDGMENTS

We thank our patients and their families for their participation in this study. We thank the Mayo Clinic Neurosurgery Clinical Research, neurosurgical residents, and operating staff for their technical and research support.

## Author contributions

Conceptualization: CRC, TCB, LPC

Sample acquisition and processing: CRC, AEW, MDH, KMA, NM, SI, KG, BTH, IJT, JJV, IFP, TCB

Data analysis: CRC, CJG, SD, TCB

Protocol and project administration: CRC, LPC, AMC, MDH, EAP, TCB

Supervision: TCB, AEW

Writing – original draft: CRC, TCB

Writing – review & editing: all authors

## Data and materials availability

All data are available as supplementary files.

## Ethics

This study was approved by the Mayo Clinic Institutional Review Board and all participants provided their consent to participate in this study. This study was performed in accordance with the Declaration of Helsinki.

## Consent

All participants have provided consent for publication.

## References

1. M. Weller, W. Wick, K. Aldape, M. Brada, M. Berger, S. M. Pfister, R. Nishikawa, M. Rosenthal, P. Y. Wen, R. Stupp, G. Reifenberger, Glioma. Nature Reviews Disease Primers 1, 15017 (2015).

2. R. Stupp, W. P. Mason, M. J. van den Bent, M. Weller, B. Fisher, M. J. Taphoorn, K. Belanger, A. A. Brandes, C. Marosi, U. Bogdahn, J. Curschmann, R. C. Janzer, S. K. Ludwin, T. Gorlia, A. Allgeier, D. Lacombe, J. G. Cairncross, E. Eisenhauer, R. O. Mirimanoff, Radiotherapy plus concomitant and adjuvant temozolomide for glioblastoma. N Engl J Med 352, 987–996 (2005).

3. M. Law, S. Yang, H. Wang, J. S. Babb, G. Johnson, S. Cha, E. A. Knopp, D. Zagzag, Glioma Grading: Sensitivity, Specificity, and Predictive Values of Perfusion MR Imaging and Proton MR Spectroscopic Imaging Compared with Conventional MR Imaging. American Journal of Neuroradiology 24, 1989–1998 (2003).

4. J. D. Bernstock, S. E. Gary, N. Klinger, P. A. Valdes, W. Ibn Essayed, H. E. Olsen, G. Chagoya, G. Elsayed, D. Yamashita, P. Schuss, F. A. Gessler, P. Paolo Peruzzi, A. K. Bag, G. K. Friedman, Standard clinical approaches and emerging modalities for glioblastoma imaging. Neuro-Oncology Advances 4, (2022).

5. P. Y. Wen, M. v. d. Bent, G. Youssef, T. F. Cloughesy, B. M. Ellingson, M. Weller, E. Galanis, D. P. Barboriak, J. d. Groot, M. R. Gilbert, R. Huang, A. B. Lassman, M. Mehta, A. M. Molinaro, M. Preusser, R. Rahman, L. K. Shankar, R. Stupp, J. E. Villanueva-Meyer, W. Wick, D. R. Macdonald, D. A. Reardon, M. A. Vogelbaum, S. M. Chang, RANO 2.0: Update to the Response Assessment in Neuro-Oncology Criteria for High- and Low-Grade Gliomas in Adults. Journal of Clinical Oncology 41, 5187–5199 (2023).

6. R. Y. Huang, M. R. Neagu, D. A. Reardon, P. Y. Wen, Pitfalls in the neuroimaging of glioblastoma in the era of antiangiogenic and immuno/targeted therapy - detecting illusive disease, defining response. Front Neurol 6, 33 (2015).

7. M. C. de Wit, H. G. de Bruin, W. Eijkenboom, P. A. Sillevis Smitt, M. J. van den Bent, Immediate post-radiotherapy changes in malignant glioma can mimic tumor progression. Neurology 63, 535–537 (2004).

8. M. Touat, A. Duran-Peña, A. Alentorn, L. Lacroix, C. Massard, A. Idbaih, Emerging circulating biomarkers in glioblastoma: promises and challenges. Expert Rev Mol Diagn 15, 1311–1323 (2015).

9. R. Soffietti, C. Bettegowda, I. K. Mellinghoff, K. E. Warren, M. S. Ahluwalia, J. F. De Groot, E. Galanis, M. R. Gilbert, K. A. Jaeckle, E. Le Rhun, R. Rudà, J. Seoane, N. Thon, Y. Umemura, M. Weller, M. J. van den Bent, M. A. Vogelbaum, S. M. Chang, P. Y. Wen, Liquid biopsy in gliomas: A RANO review and proposals for clinical applications. Neuro-Oncology 24, 855–871 (2022).

10. S. J. Bagley, S. A. Nabavizadeh, J. J. Mays, J. E. Till, J. B. Ware, S. Levy, W. Sarchiapone, J. Hussain, T. Prior, S. Guiry, Clinical utility of plasma cell-free DNA in adult patients with newly diagnosed glioblastoma: a pilot prospective study. Clinical Cancer Research 26, 397–407 (2020).

11. A. M. Miller, R. H. Shah, E. I. Pentsova, M. Pourmaleki, S. Briggs, N. Distefano, Y. Zheng, A. Skakodub, S. A. Mehta, C. Campos, W.-Y. Hsieh, S. D. Selcuklu, L. Ling, F. Meng, X. Jing, A. Samoila, T. A. Bale, D. W. Y. Tsui, C. Grommes, A. Viale, M. M. Souweidane, V. Tabar, C. W. Brennan, A. S. Reiner, M. Rosenblum, K. S. Panageas, L. M. DeAngelis, R. J. Young, M. F. Berger, I. K. Mellinghoff, Tracking tumour evolution in glioma through liquid biopsies of cerebrospinal fluid. Nature 565, 654–658 (2019).

12. E. Le Rhun, J. Seoane, M. Salzet, R. Soffietti, M. Weller, Liquid biopsies for diagnosing and monitoring primary tumors of the central nervous system. Cancer Lett 480, 24–28 (2020).

13. J. F. Deeken, W. Löscher, The blood-brain barrier and cancer: transporters, treatment, and Trojan horses. Clin Cancer Res 13, 1663–1674 (2007).

14. J. Rincon-Torroella, H. Khela, A. Bettegowda, C. Bettegowda, Biomarkers and focused ultrasound: the future of liquid biopsy for brain tumor patients. J Neurooncol 156, 33–48 (2022).

15. Y. Meng, C. B. Pople, S. Suppiah, M. Llinas, Y. Huang, A. Sahgal, J. Perry, J. Keith, B. Davidson, C. Hamani, Y. Amemiya, A. Seth, H. Leong, C. C. Heyn, I. Aubert, K. Hynynen, N. Lipsman, MR-guided focused ultrasound liquid biopsy enriches circulating biomarkers in patients with brain tumors. Neuro Oncol 23, 1789–1797 (2021).

16. J. Yuan, L. Xu, C.-Y. Chien, Y. Yang, Y. Yue, S. Fadera, A. H. Stark, K. E. Schwetye, A. Nazeri, R. Desai, U. Athiraman, A. A. Chaudhuri, H. Chen, E. C. Leuthardt, First-in-human prospective trial of sonobiopsy in high-grade glioma patients using neuronavigation-guided focused ultrasound. npj Precision Oncology 7, 92 (2023).

17. C. Bettegowda, M. Sausen, R. J. Leary, I. Kinde, Y. Wang, N. Agrawal, B. R. Bartlett, H. Wang, B. Luber, R. M. Alani, E. S. Antonarakis, N. S. Azad, A. Bardelli, H. Brem, J. L. Cameron, C. C. Lee, L. A. Fecher, G. L. Gallia, P. Gibbs, D. Le, R. L. Giuntoli, M. Goggins, M. D. Hogarty, M. Holdhoff, S. M. Hong, Y. Jiao, H. H. Juhl, J. J. Kim, G. Siravegna, D. A. Laheru, C. Lauricella, M. Lim, E. J. Lipson, S. K. Marie, G. J. Netto, K. S. Oliner, A. Olivi, L. Olsson, G. J. Riggins, A. Sartore-Bianchi, K. Schmidt, M. Shih l, S. M. Oba-Shinjo, S. Siena, D. Theodorescu, J. Tie, T. T. Harkins, S. Veronese, T. L. Wang, J. D. Weingart, C. L. Wolfgang, L. D. Wood, D. Xing, R. H. Hruban, J. Wu, P. J. Allen, C. M. Schmidt, M. A. Choti, V. E. Velculescu, K. W. Kinzler, B. Vogelstein, N. Papadopoulos, L. A. Diaz, Jr., Detection of circulating tumor DNA in early- and late-stage human malignancies. Sci Transl Med 6, 224ra224 (2014).

18. L. Zhu, G. Cheng, D. Ye, A. Nazeri, Y. Yue, W. Liu, X. Wang, G. P. Dunn, A. A. Petti, E. C. Leuthardt, H. Chen, Focused Ultrasound-enabled Brain Tumor Liquid Biopsy. Sci Rep 8, 6553 (2018).

19. F. Xiao, S. Lv, Z. Zong, L. Wu, X. Tang, W. Kuang, P. Zhang, X. Li, J. Fu, M. Xiao, M. Wu, L. Wu, X. Zhu, K. Huang, H. Guo, Cerebrospinal fluid biomarkers for brain tumor detection: clinical roles and current progress. Am J Transl Res 12, 1379–1396 (2020).

20. L. Escudero, A. Llort, A. Arias, A. Diaz-Navarro, F. Martínez-Ricarte, C. Rubio-Perez, R. Mayor, G. Caratù, E. Martínez-Sáez, É. Vázquez-Méndez, I. Lesende-Rodríguez, R. Hladun, L. Gros, S. Ramón y Cajal, M. A. Poca, X. S. Puente, J. Sahuquillo, S. Gallego, J. Seoane, Circulating tumour DNA from the cerebrospinal fluid allows the characterisation and monitoring of medulloblastoma. Nature Communications 11, 5376 (2020).

21. J. M. Kros, D. M. Mustafa, L. J. M. Dekker, P. A. E. Sillevis Smitt, T. M. Luider, P.-P. Zheng, Circulating glioma biomarkers. Neuro-Oncology 17, 343–360 (2014).

22. A. P. Y. Liu, K. S. Smith, R. Kumar, L. Paul, L. Bihannic, T. Lin, K. K. Maass, K. W. Pajtler, M. Chintagumpala, J. M. Su, E. Bouffet, M. J. Fisher, S. Gururangan, R. Cohn, T. Hassall, J. R. Hansford, P. Klimo, Jr., F. A. Boop, C. F. Stewart, J. H. Harreld, T. E. Merchant, R. G. Tatevossian, G. Neale, M. Lear, J. M. Klco, B. A. Orr, D. W. Ellison, R. J. Gilbertson, A. Onar-Thomas, A. Gajjar, G. W. Robinson, P. A. Northcott, Serial assessment of measurable residual disease in medulloblastoma liquid biopsies. Cancer Cell 39, 1519–1530.e1514 (2021).

23. C. Riviere-Cazaux, X. Dong, W. Mo, C. Dai, L. P. Carlstrom, A. Munoz-Casabella, R. Kumar, K. Ghadimi, C. L. Nesvick, K. M. Andersen, M. D. Hoplin, N. Canaday, I. Jusue-Torres, N. Malik, J. L. Campian, M. W. Ruff, J. H. Uhm, J. E. E. Passow, T. J. Kaufmann, D. M. Routman, S. H. Kizilbash, A. E. Warrington, R. B. Jenkins, P. Du, S. Jia, T. C. Burns, Longitudinal glioma monitoring via cerebrospinal fluid cell-free DNA: one patient at a time. medRxiv, 2024.2002.2021.24303164 (2024).

24. A. E. McEwen, S. E. S. Leary, C. M. Lockwood, Beyond the Blood: CSF-Derived cfDNA for Diagnosis and Characterization of CNS Tumors. Front Cell Dev Biol 8, 45 (2020).

25. M. D. White, R. H. Klein, B. Shaw, A. Kim, M. Subramanian, J. L. Mora, A. Giobbie-Hurder, D. Nagabhushan, A. Jain, M. Singh, B. M. Kuter, N. Nayyar, M. S. Bertalan, J. H. Stocking, S. C. Markson, M. Lastrapes, C. Alvarez-Breckenridge, D. P. Cahill, G. Gydush, J. Rhoades, D. Rotem, V. A. Adalsteinsson, M. Mahar, A. Kaplan, K. Oh, R. J. Sullivan, E. Gerstner, S. L. Carter, P. K. Brastianos, Detection of Leptomeningeal Disease Using Cell-Free DNA From Cerebrospinal Fluid. JAMA Network Open 4, e2120040–e2120040 (2021).

26. R. Mair, F. Mouliere, Cell-free DNA technologies for the analysis of brain cancer. British Journal of Cancer 126, 371–378 (2022).

27. C. Riviere-Cazaux, J. M. Lacey, L. P. Carlstrom, W. J. Laxen, A. Munoz-Casabella, M. D. Hoplin, S. Ikram, A. B. Zubair, K. M. Andersen, A. E. Warrington, P. A. Decker, T. J. Kaufmann, J. L. Campian, J. E. Eckel-Passow, S. H. Kizilbash, S. Tortorelli, T. C. Burns, Cerebrospinal fluid 2-hydroxyglutarate as a monitoring biomarker for IDH-mutant gliomas. Neuro-Oncology Advances 5, (2023).

28. Y. Fujita, L. Nunez-Rubiano, A. Dono, A. Bellman, M. Shah, J. C. Rodriguez, V. Putluri, A. H. M. Kamal, N. Putluri, R. F. Riascos, J. J. Zhu, Y. Esquenazi, L. Y. Ballester, IDH1 p.R132H ctDNA and D-2-hydroxyglutarate as CSF biomarkers in patients with IDH-mutant gliomas. J Neurooncol 159, 261–270 (2022).

29. L. Y. Ballester, G. Lu, S. Zorofchian, V. Vantaku, V. Putluri, Y. Yan, O. Arevalo, P. Zhu, R. F. Riascos, A. Sreekumar, Y. Esquenazi, N. Putluri, J.-J. Zhu, Analysis of cerebrospinal fluid metabolites in patients with primary or metastatic central nervous system tumors. Acta Neuropathologica Communications 6, 85 (2018).

30. J. Kalinina, J. Ahn, N. S. Devi, L. Wang, Y. Li, J. J. Olson, M. Glantz, T. Smith, E. L. Kim, A. Giese, R. L. Jensen, C. C. Chen, B. S. Carter, H. Mao, M. He, E. G. Van Meir, Selective Detection of the D-enantiomer of 2-Hydroxyglutarate in the CSF of Glioma Patients with Mutated Isocitrate Dehydrogenase. Clin Cancer Res 22, 6256–6265 (2016).

31. J. Kalinina, J. Peng, J. C. Ritchie, E. G. Van Meir, Proteomics of gliomas: initial biomarker discovery and evolution of technology. Neuro Oncol 13, 926–942 (2011).

32. M. U. Schuhmann, H. D. Zucht, R. Nassimi, G. Heine, C. G. Schneekloth, H. J. Stuerenburg, H. Selle, Peptide screening of cerebrospinal fluid in patients with glioblastoma multiforme. Eur J Surg Oncol 36, 201–207 (2010).

33. D. Schmid, U. Warnken, P. Latzer, D. C. Hoffmann, J. Roth, S. Kutschmann, H. Jaschonek, P. Rübmann, M. Foltyn, P. Vollmuth, F. Winkler, C. Seliger, M. Felix, F. Sahm, J. Haas, D. Reuss, M. Bendszus, B. Wildemann, A. von Deimling, W. Wick, T. Kessler, Diagnostic biomarkers from proteomic characterization of cerebrospinal fluid in patients with brain malignancies. J Neurochem 158, 522–538 (2021).

34. F. W. Khwaja, M. S. Reed, J. J. Olson, B. J. Schmotzer, G. Y. Gillespie, A. Guha, M. D. Groves, S. Kesari, J. Pohl, E. G. Van Meir, Proteomic identification of biomarkers in the cerebrospinal fluid (CSF) of astrocytoma patients. J Proteome Res 6, 559–570 (2007).

35. F. W. Khwaja, J. S. Duke-Cohan, D. J. Brat, E. G. Van Meir, Attractin is elevated in the cerebrospinal fluid of patients with malignant astrocytoma and mediates glioma cell migration. Clin Cancer Res 12, 6331–6336 (2006).

36. N. Mikolajewicz, S. Khan, M. Trifoi, A. Skakdoub, V. Ignatchenko, S. Mansouri, J. Zuccato, B. E. Zacharia, M. Glantz, G. Zadeh, J. Moffat, T. Kislinger, A. Mansouri, Leveraging the CSF proteome toward minimally-invasive diagnostics surveillance of brain malignancies. Neuro-Oncology Advances 4, (2022).

37. N. Rostgaard, M. H. Olsen, M. Ottenheijm, L. Drici, A. H. Simonsen, P. Plomgaard, H. Gredal, H. H. Poulsen, H. Zetterberg, K. Blennow, S. G. Hasselbalch, N. MacAulay, M. Juhler, Differential proteomic profile of lumbar and ventricular cerebrospinal fluid. Fluids Barriers CNS 20, 6 (2023).

38. Y. Kim, J. Jeon, S. Mejia, C. Q. Yao, V. Ignatchenko, J. O. Nyalwidhe, A. O. Gramolini, R. S. Lance, D. A. Troyer, R. R. Drake, P. C. Boutros, O. J. Semmes, T. Kislinger, Targeted proteomics identifies liquid-biopsy signatures for extracapsular prostate cancer. Nat Commun 7, 11906 (2016).

39. U. Kuzmanov, N. Musrap, H. Kosanam, C. R. Smith, I. Batruch, A. Dimitromanolakis, E. P. Diamandis, Glycoproteomic identification of potential glycoprotein biomarkers in ovarian cancer proximal fluids. Clin Chem Lab Med 51, 1467–1476 (2013).

40. L. Gold, D. Ayers, J. Bertino, C. Bock, A. Bock, E. N. Brody, J. Carter, A. B. Dalby, B. E. Eaton, T. Fitzwater, D. Flather, A. Forbes, T. Foreman, C. Fowler, B. Gawande, M. Goss, M. Gunn, S. Gupta, D. Halladay, J. Heil, J. Heilig, B. Hicke, G. Husar, N. Janjic, T. Jarvis, S. Jennings, E. Katilius, T. R. Keeney, N. Kim, T. H. Koch, S. Kraemer, L. Kroiss, N. Le, D. Levine, W. Lindsey, B. Lollo, W. Mayfield, M. Mehan, R. Mehler, S. K. Nelson, M. Nelson, D. Nieuwlandt, M. Nikrad, U. Ochsner, R. M. Ostroff, M. Otis, T. Parker, S. Pietrasiewicz, D. I. Resnicow, J. Rohloff, G. Sanders, S. Sattin, D. Schneider, B. Singer, M. Stanton, A. Sterkel, A. Stewart, S. Stratford, J. D. Vaught, M. Vrkljan, J. J. Walker, M. Watrobka, S. Waugh, A. Weiss, S. K. Wilcox, A. Wolfson, S. K. Wolk, C. Zhang, D. Zichi, Aptamer-based multiplexed proteomic technology for biomarker discovery. PLoS One 5, e15004 (2010).

41. J. C. Rohloff, A. D. Gelinas, T. C. Jarvis, U. A. Ochsner, D. J. Schneider, L. Gold, N. Janjic, Nucleic Acid Ligands With Protein-like Side Chains: Modified Aptamers and Their Use as Diagnostic and Therapeutic Agents. Molecular Therapy - Nucleic Acids 3, e201 (2014).

42. P. Halleux, S. Schurmans, S. N. Schiffman, R. Lecocq, J. L. Conreur, J. Dumont, J. J. Vanderhaeghen, Calcium binding protein calcyphosine in dog central astrocytes and ependymal cells and in peripheral neurons. J Chem Neuroanat 15, 239–250 (1998).

43. V. Ribes, J. Briscoe, Establishing and interpreting graded Sonic Hedgehog signaling during vertebrate neural tube patterning: the role of negative feedback. Cold Spring Harb Perspect Biol 1, a002014 (2009).

44. C. Riviere-Cazaux, L. P. Carlstrom, K. Rajani, A. Munoz-Casabella, M. Rahman, A. Gharibi-Loron, D. A. Brown, K. J. Miller, J. J. White, B. T. Himes, I. Jusue-Torres, S. Ikram, S. C. Ransom, R. Hirte, J.-H. Oh, W. F. Elmquist, J. N. Sarkaria, R. A. Vaubel, M. Rodriguez, A. E. Warrington, S. H. Kizilbash, T. C. Burns, Blood-brain barrier disruption defines the extracellular metabolome of live human high-grade gliomas. Communications Biology 6, 653 (2023).

45. A. T. Argaw, B. T. Gurfein, Y. Zhang, A. Zameer, G. R. John, VEGF-mediated disruption of endothelial CLN-5 promotes blood-brain barrier breakdown. Proc Natl Acad Sci U S A 106, 1977–1982 (2009).

46. M. R. Gilbert, J. J. Dignam, T. S. Armstrong, J. S. Wefel, D. T. Blumenthal, M. A. Vogelbaum, H. Colman, A. Chakravarti, S. Pugh, M. Won, R. Jeraj, P. D. Brown, K. A. Jaeckle, D. Schiff, V. W. Stieber, D. G. Brachman, M. Werner-Wasik, I. W. Tremont-Lukats, E. P. Sulman, K. D. Aldape, W. J. Curran, M. P. Mehta, A Randomized Trial of Bevacizumab for Newly Diagnosed Glioblastoma. New England Journal of Medicine 370, 699–708 (2014).

47. R. Radu, G. E. D. Petrescu, R. M. Gorgan, F. M. Brehar, GFAPδ: A Promising Biomarker and Therapeutic Target in Glioblastoma. Front Oncol 12, 859247 (2022).

48. C. S. Jung, C. Foerch, A. Schänzer, A. Heck, K. H. Plate, V. Seifert, H. Steinmetz, A. Raabe, M. Sitzer, Serum GFAP is a diagnostic marker for glioblastoma multiforme. Brain 130, 3336–3341 (2007).

49. J. Gállego Pérez-Larraya, S. Paris, A. Idbaih, C. Dehais, F. Laigle-Donadey, S. Navarro, L. Capelle, K. Mokhtari, Y. Marie, M. Sanson, Diagnostic and prognostic value of preoperative combined GFAP, IGFBP-2, and YKL-40 plasma levels in patients with glioblastoma. Cancer 120, 3972–3980 (2014).

50. M. Osada, H. L. Park, M. J. Park, J. W. Liu, G. Wu, B. Trink, D. Sidransky, A p53-type response element in the GDF15 promoter confers high specificity for p53 activation. Biochem Biophys Res Commun 354, 913–918 (2007).

51. M. V. Goldberg, C. G. Drake, LAG-3 in Cancer Immunotherapy. Curr Top Microbiol Immunol 344, 269–278 (2011).

52. K. Singh, K. M. Hotchkiss, I. F. Parney, J. De Groot, S. Sahebjam, N. Sanai, M. Platten, E. Galanis, M. Lim, P. Y. Wen, G. Minniti, H. Colman, T. F. Cloughesy, M. P. Mehta, M. Geurts, I. Arrillaga-Romany, A. Desjardins, K. Tanner, S. Short, D. Arons, E. Duke, W. Wick, S. J. Bagley, D. M. Ashley, P. Kumthekar, R. Verhaak, A. J. Chalmers, A. P. Patel, C. Watts, P. E. Fecci, T. T. Batchelor, M. Weller, M. A. Vogelbaum, M. Preusser, M. S. Berger, M. Khasraw, Correcting the drug development paradigm for glioblastoma requires serial tissue sampling. Nature Medicine 29, 2402–2405 (2023).

53. H. Chen, J. Ji, L. Zhang, T. Chen, Y. Zhang, F. Zhang, J. Wang, Y. Ke, Inflammatory responsive neutrophil-like membrane-based drug delivery system for post-surgical glioblastoma therapy. J Control Release 362, 479–488 (2023).

54. A. M. Knudsen, B. Halle, O. Cédile, M. Burton, C. Baun, H. Thisgaard, A. Anand, C. Hubert, M. Thomassen, S. R. Michaelsen, B. B. Olsen, R. H. Dahlrot, R. Bjerkvig, J. D. Lathia, B. W. Kristensen, Surgical resection of glioblastomas induces pleiotrophin-mediated self-renewal of glioblastoma stem cells in recurrent tumors. Neuro Oncol 24, 1074–1087 (2022).

55. E. Aasebø, J. A. Opsahl, Y. Bjørlykke, K. M. Myhr, A. C. Kroksveen, F. S. Berven, Effects of blood contamination and the rostro-caudal gradient on the human cerebrospinal fluid proteome. PLoS One 9, e90429 (2014).

56. G. H. Sato, The role of serum in cell culture. Biochemical actions of hormones 3, 391–396 (1975).

57. Y. Wang, S. Springer, M. Zhang, K. W. McMahon, I. Kinde, L. Dobbyn, J. Ptak, H. Brem, K. Chaichana, G. L. Gallia, Z. L. Gokaslan, M. L. Groves, G. I. Jallo, M. Lim, A. Olivi, A. Quinones-Hinojosa, D. Rigamonti, G. J. Riggins, D. M. Sciubba, J. D. Weingart, J.-P. Wolinsky, X. Ye, S. M. Oba-Shinjo, S. K. N. Marie, M. Holdhoff, N. Agrawal, L. A. Diaz, N. Papadopoulos, K. W. Kinzler, B. Vogelstein, C. Bettegowda, Detection of tumor-derived DNA in cerebrospinal fluid of patients with primary tumors of the brain and spinal cord. Proceedings of the National Academy of Sciences 112, 9704–9709 (2015).

58. L. S. Graham, C. C. Pritchard, M. T. Schweizer, Hypermutation, Mismatch Repair Deficiency, and Defining Predictors of Response to Checkpoint Blockade. Clin Cancer Res 27, 6662–6665 (2021).

59. S. Harris-Bookman, D. Mathios, A. M. Martin, Y. Xia, E. Kim, H. Xu, Z. Belcaid, M. Polanczyk, T. Barberi, D. Theodros, J. Kim, J. M. Taube, P. C. Burger, M. Selby, C. Taitt, A. Korman, X. Ye, C. G. Drake, H. Brem, D. M. Pardoll, M. Lim, Expression of LAG-3 and efficacy of combination treatment with anti-LAG-3 and anti-PD-1 monoclonal antibodies in glioblastoma. Int J Cancer 143, 3201–3208 (2018).

60. J. Timsina, D. Gomez-Fonseca, L. Wang, A. Do, D. Western, I. Alvarez, M. Aguilar, P. Pastor, R. L. Henson, E. Herries, C. Xiong, S. E. Schindler, A. M. Fagan, R. J. Bateman, M. Farlow, J. C. Morris, R. J. Perrin, K. Moulder, J. Hassenstab, J. Vöglein, J. Chhatwal, H. Mori, Y. J. Sung, C. Cruchaga, Comparative Analysis of Alzheimer’s Disease Cerebrospinal Fluid Biomarkers Measurement by Multiplex SOMAscan Platform and Immunoassay-Based Approach. J Alzheimers Dis 89, 193–207 (2022).

61. M. D. Robinson, A. Oshlack, A scaling normalization method for differential expression analysis of RNA-seq data. Genome biology 11, 1–9 (2010).

