## Supplementary Figures for "The dynamic impact of location and resection on the glioma CSF proteome"

**Supplementary Materials**

**Supplementary Figure 1. Enrichment analyses with paired pre-versus-early post-resection CSF samples from patients with gliomas.**

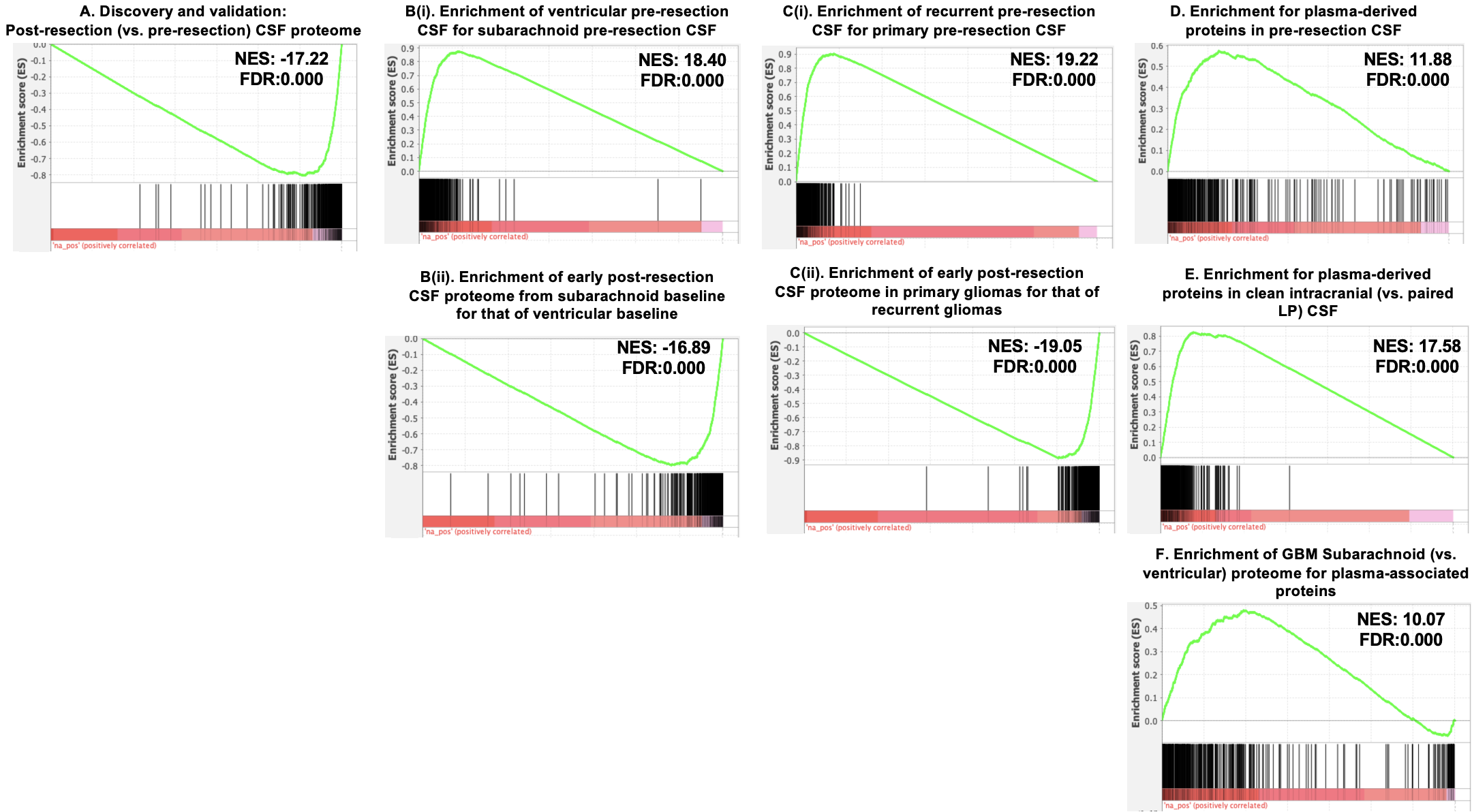

**A.** Enrichment of the discovery cohort’s pre-versus-early post resection CSF proteome for the validation cohort’s early post-resection CSF proteome, indicating overlap in the early post-resection CSF proteomes across discovery and validation cohorts.

**B**. The 20 paired pre-versus-post-resection CSF samples were split into two cohorts based on whether the pre-resection CSF sample was obtained from the subarachnoid or ventricular space. The ventricular cohort was used for the protein library sets (top 350 proteins from fold-change). The subarachnoid cohort was used for the ranked list (pre-versus-early post-resection). Enrichment analysis was performed, demonstrating **(i)** enrichment of the pre-resection ventricular and subarachnoid CSF proteomes for one another, when each were compared to post-resection CSF and **(ii)** enrichment of the early post-resection CSF proteomes for both the ventricular and subarachnoid cohorts.

**C**. Similar analyses to Supp. Figure 1B were performed, splitting the cohorts based on whether the patient had a primary or recurrent glioma. Enrichment analysis revealed **(i)** enrichment of the pre-resection CSF proteome of primary and recurrent glioma for one another, when each were compared to their respective early post-resection CSF samples, and **(ii)** enrichment of the early post-resection CSF proteomes for both the primary and recurrent cohorts.

**D-F.** Enrichment analysis for plasma-derived proteins, as defined based on paired bloody and clean CSF samples (top 350 proteins from fold-change), was evaluated in the **D)** pre-versus-early post-resection ranked protein list (n=20 pairs), **E)** intracranial versus lumbar puncture ranked protein list (n=14 pairs), and **F)** glioblastoma (GBM) subarachnoid versus ventricular ranked protein list (unpaired; n=23 subarachnoid versus n=20 ventricular).

NES= normalized enrichment score. FDR = false discovery rate.

**Supplementary Figure 2. Enrichment analyses in 9 paired early and delayed post-resection CSF samples from patients with gliomas.**

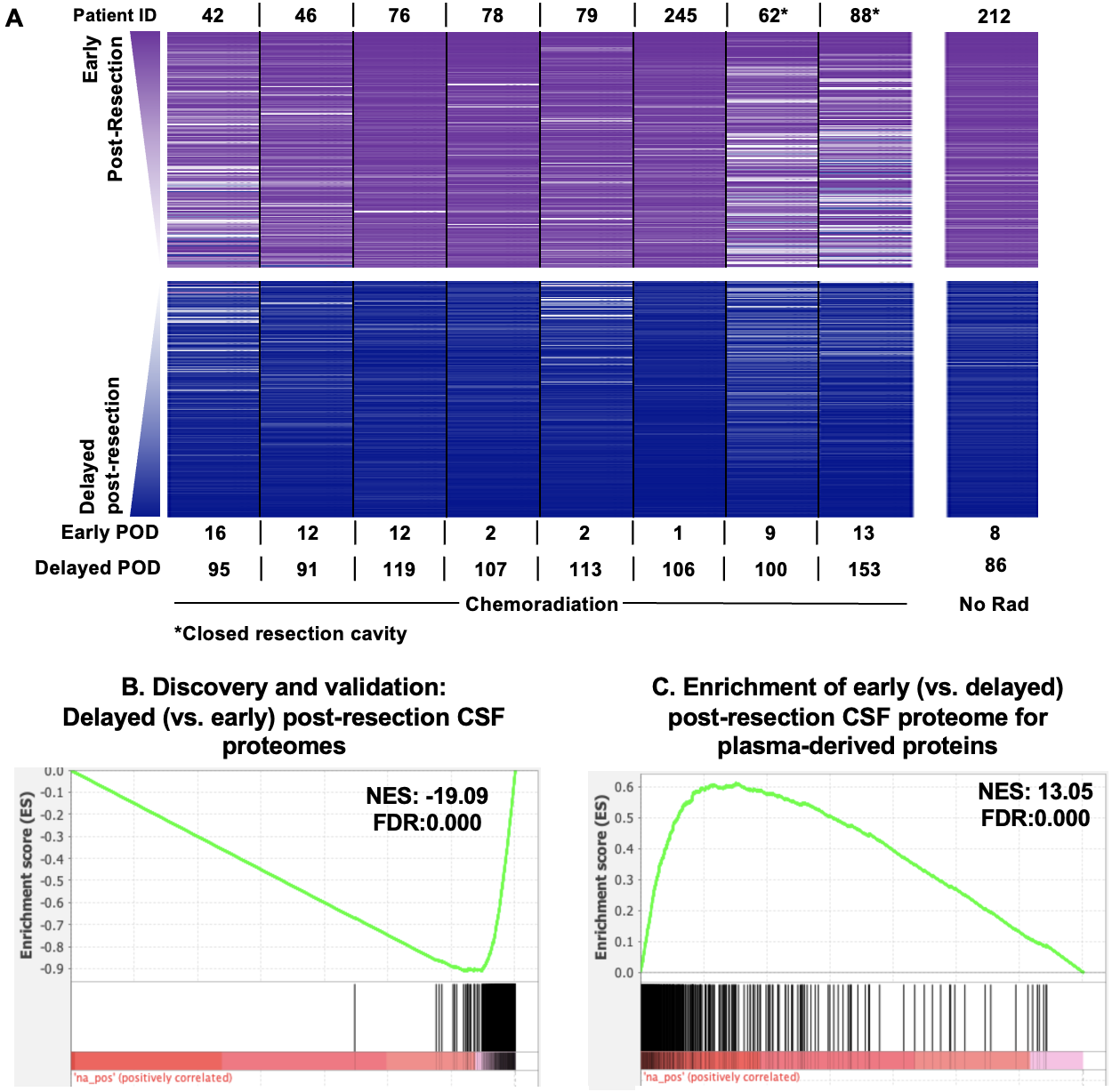

**A.** Ranked fold-change lists were generated from each patient’s paired early-versus-delayed post-resection CSF samples (n=9), as in Figure 3Bi. Ranked protein order is shown as a heatmap from 1 (higher early post-resection; purple) to 7,011 (higher delayed post-resection; blue). Eight patients received chemoradiation during this time frame. One patient (#212) did not. The average rank across the 8 patients who received chemoradiation was used to rank all 9 ranked lists. The top and bottom 200 proteins are shown.

**B.** Enrichment in the discovery cohort’s early-versus-delayed post-resection CSF proteome for the validation cohort’s delayed post-resection CSF proteome, indicating overlap in the delayed post-resection CSF proteomes across discovery and validation cohorts.

**C**. Enrichment analysis for plasma-derived proteins, as defined based on paired bloody and clean CSF samples (top 350 proteins from fold-change), was evaluated in the early-versus-delayed post-resection ranked protein list.

**Supplementary Figure 3. Enrichment analyses in the 11 paired pre-resection versus delayed post-resection CSF samples from patients with gliomas.**

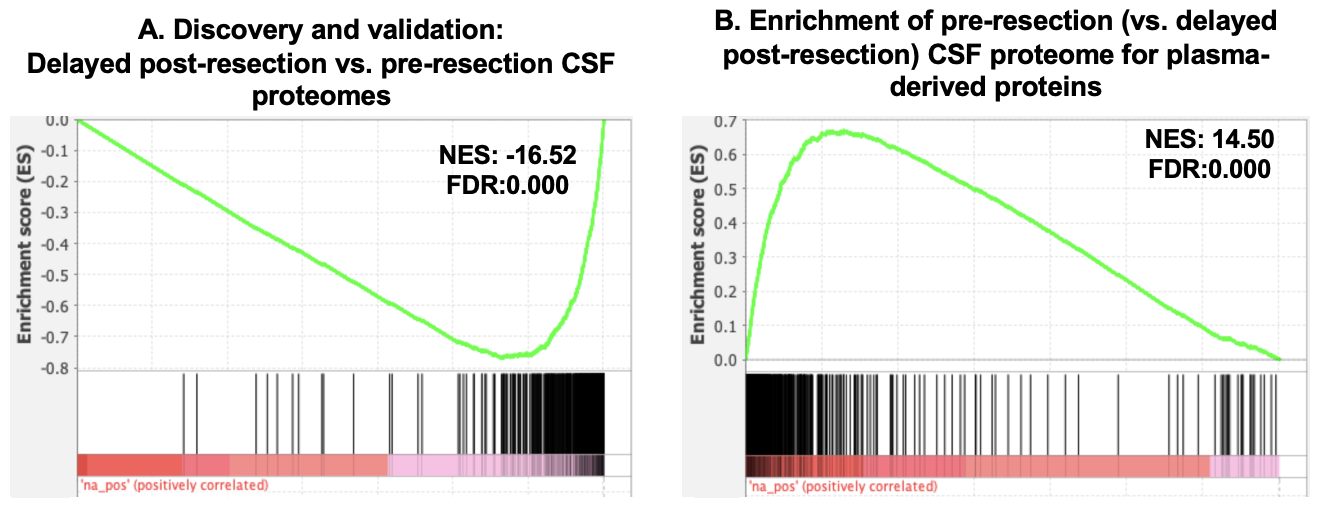

**A.** Enrichment in the discovery cohort’s pre-resection-versus-delayed post-resection CSF proteome for the validation cohort’s delayed post-resection CSF proteome, indicating overlap in the delayed post-resection CSF proteomes across discovery and validation cohorts when compared to the pre-resection CSF proteome.

**B**. Enrichment analysis for plasma-derived proteins, as defined based on paired bloody and clean CSF samples (top 350 proteins from fold-change), was evaluated in the pre-resection-versus-delayed post-resection ranked protein list.

**Supplementary Figure 4**. **Evaluation of candidate monitoring and pharmacodynamic proteins in longitudinal CSF samples.**

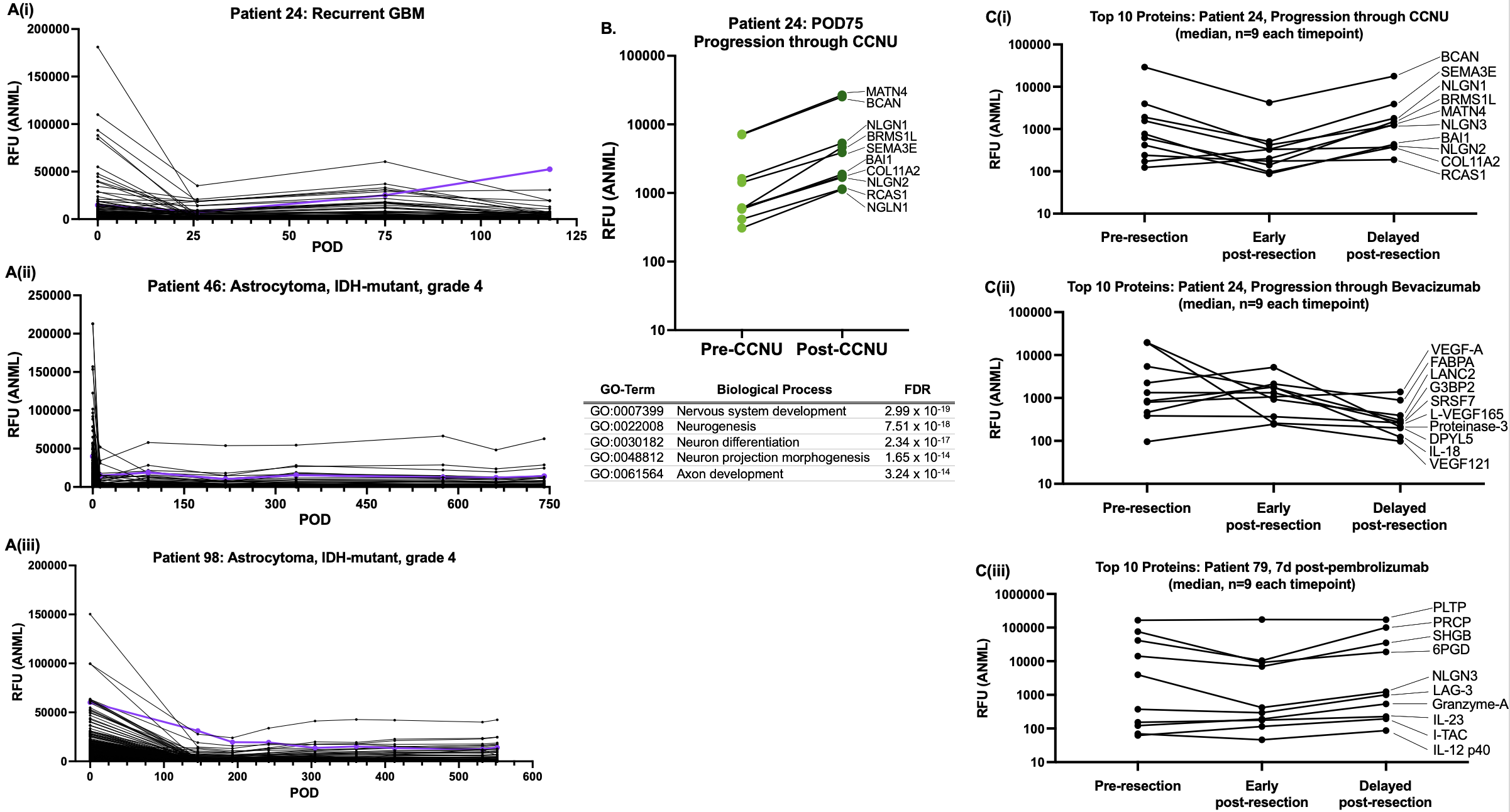

**A.** The top 25% of proteins decreasing with resection from **Fig. 2A** were plotted over time starting from resection (post-operative day (POD) 0) in patients **(i)** 24, **(ii)** 46, and **(iii)** 98. Brevican (BCAN) is plotted in purple as the identified protein that best correlated with disease course across all three patients.

**B.** Protein fold-changes were calculated in the samples obtained after versus before progression through CCNU (or lomustine) in patient 24. The top 10 proteins based on fold-change were plotted. Protein network enrichment analysis was performed on the top 200 proteins based on fold-change of proteins pre-versus-post-progression through CCNU – five pathways are shown on the table.

**C.** Nine patients had CSF samples obtained at pre-resection, early post-resection, and delayed post-resection timepoints. The median abundance for each of the 10 proteins shown in Supplementary Figure 4B and Figure 4Di-ii are plotted from each timepoint in **(i), (ii),** and **(iii)** respectively. The median is based on 9 patients at each timepoint.
